# Genetic and environmental determinants of diastolic heart function

**DOI:** 10.1101/2021.06.07.21257302

**Authors:** Marjola Thanaj, Johanna Mielke, Kathryn A. McGurk, Wenjia Bai, Nicoló Savioli, Antonio de Marvao, Hannah V. Meyer, Lingyao Zeng, Florian Sohler, Martin R. Wilkins, James S. Ware, Christian Bender, Daniel Rueckert, Aidan MacNamara, Daniel F. Freitag, Declan P. O’Regan

## Abstract

Diastole is the sequence of physiological events that occur in the heart during ventricular filling and principally depends on myocardial relaxation and chamber stiffness. Abnormal diastolic function is related to many cardiovascular disease processes and is predictive of health outcomes, but its genetic architecture is largely unknown. Here, we use machine learning cardiac motion analysis to measure diastolic functional traits in 39,559 participants of UK Biobank and perform a genome-wide association study. We identified 9 significant, independent loci near genes that are associated with maintaining sarcomeric function under biomechanical stress and genes implicated in the development of cardiomyopathy. Age, sex and diabetes were independent predictors of diastolic function and we found a causal relationship between ventricular stiffness and heart failure. Our results provide novel insights into the genetic and environmental factors influencing diastolic function that are relevant for identifying causal relationships and tractable targets in heart failure.

## Main

Diastole is not a passive phase of the cardiac cycle, but is a complex sequence of inter-related physiological processes dependent on myocardial relaxation, stiffness and recoil, that are modulated by loading conditions, heart rate, and contractile function^1,2^. Diastolic function therefore plays a central role in determining left ventricular filling and stroke volume with dysfunction shown to be a predictor of major adverse cardiovascular events and all cause mortality^3^. Decline in diastolic function is also a hallmark of cardiac ageing which occurs through multiple pro-fibrotic and energetic pathways^4,5^. While several candidate genes have been implicated in various systolic function phenotypes through genome wide association studies (GWAS)^6,7^, the genetic architecture of diastolic function and causal associations with disease are largely unknown. Efforts to better define the molecular mechanisms of diastolic dysfunction could enable the development of innovative therapies for many cardiovascular disease states^8^.

Pre-clinical models of diastolic dysfunction are associated with alterations in left ventricular stiffness on atomic force microscopy that occur at the level of the cardiomyocyte sarcomere as well as due to extracellular matrix protein expansion^9^. Such tissue level changes can be assessed at macroscopic scale in human populations through analysis of diastolic mechanics. Here we use data from participants in UK Biobank with cardiac magnetic resonance imaging (CMR)^10^, and apply deep learning computer vision techniques for precision motion analysis to derive image-based phenotypes of diastolic function^11,12^. In a GWAS of diastolic traits we identify associated loci that map to genes involved in actin assembly, cardiac myocyte survival, and heart failure phenotypes. We also describe the relationship between diastolic function and cardiovascular risk factors, and identify potential causal relationships with disease through Mendelian randomisation.

## Results

### Study Overview

We analysed CMR data from 39,559 participants in UK Biobank using machine learning segmentation and motion tracking to measure three validated parameters of diastolic function - radial and longitudinal peak early diastolic strain rate (PDSR_*rr*_ and PDSR_*ll*_) (Figure 1), and maximum body surface area-indexed left atrial volume (LAVmax_*i*_)^13^. A flow chart of the analysis steps is depicted in Extended Data Fig 1. Baseline characteristics of the population are shown in Extended Data Table 1. The population was partitioned into discovery and validation sets by pre-defined criteria. To assess the association between these diastolic function traits and other clinical measurements, we further considered a broad selection of 30 imaging and 110 non-imaging phenotypes that included biophysical data and circulating biomarkers (Supplementary Data 1). Independent GWASs were undertaken for each image-derived phenotype and heritability estimated. We used a phenome-wide association study (PheWAS) to identify multiple phenotypes associated with a polygenic risk score (PRS) for diastolic function. Potentially causal associations were examined using 2-sample Mendelian randomisation (MR). The results are reported in accordance with GWAS reporting guidelines and a checklist is provided as Supplementary Information^14^.

**Figure 1.**
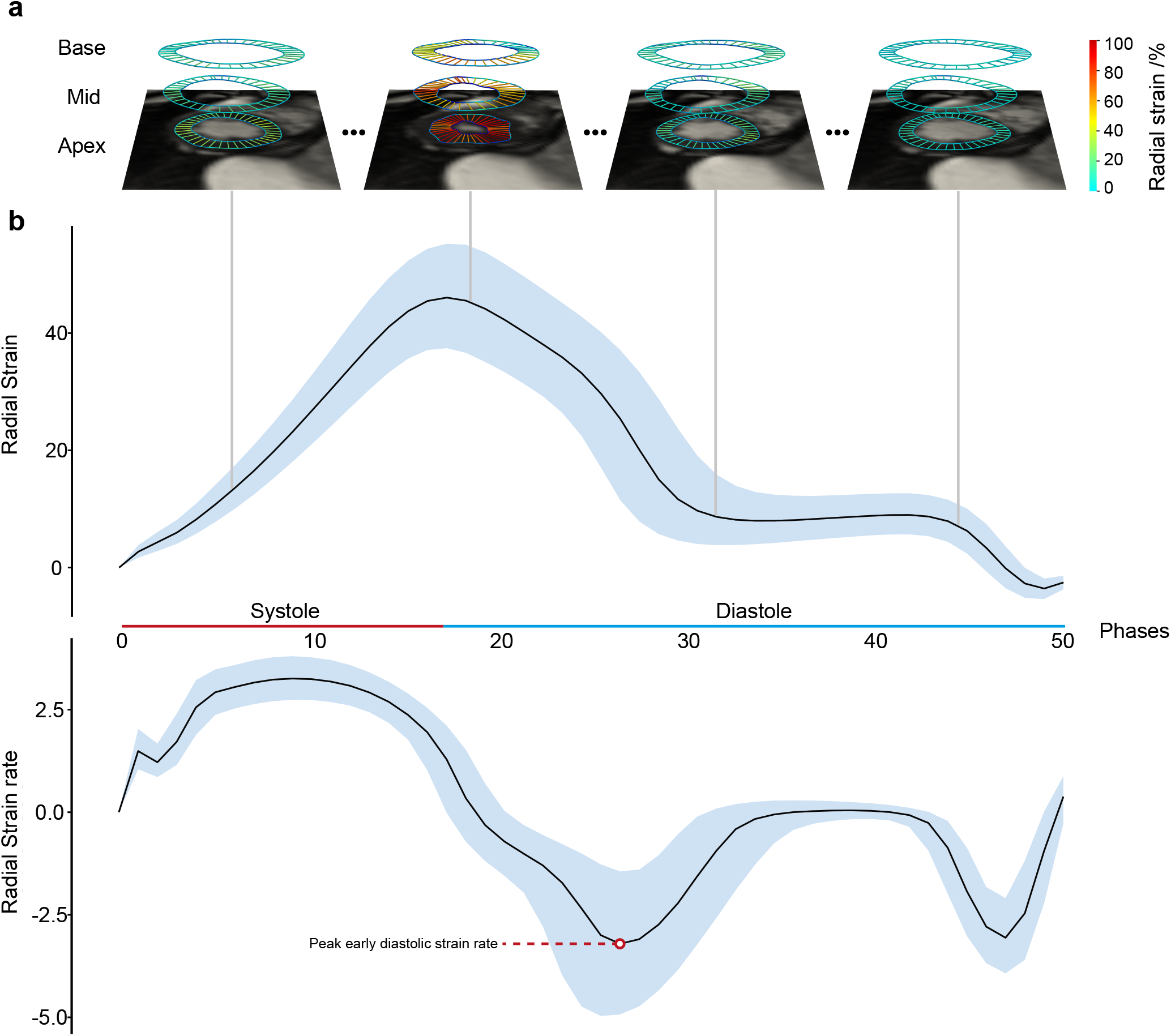
Analysis of cardiac motion. Motion analysis of cardiac magnetic resonance imaging performed on left ventricular short axis cines. A) An example from one individual where deep learning segmentation and image registration were used to determine the radial components of myocardial deformation. Data from the basal, mid-ventricular, and apical levels are shown at four representative phases from the 50 acquired. B) Radial strain and strain rate (first derivative of strain) for all UK Biobank subjects (median and interquartile ranges, n=39,559 individuals).

### Imaging and non-imaging phenotype associations

Strain rates declined with age and were lower in men (*P <* 10^−16^ for both associations) (Figure 2), but no univariable association was observed between age and LAVmax_*i*_ (Extended Data Fig 2). Multiple linear regression analysis was used to develop a model for predicting each diastolic trait from demographic, haemodynamic and cardiovascular risk factors (Figure 3a, Extended Data Fig 3). In this multivariable analysis strain rate and left atrial volumes were negatively associated with age, sex and pulse rate in the full model (*P* < 10^−16^ for all associations). Significant associations were also observed for body surface area (BSA) and systolic blood pressure (SBP). Diabetes also added significantly to the associations with the diastolic function traits in the model (PDSR_*ll*_: *P* = 2.36 × 10^*−*8^; PDSR_*rr*_: *P* = 9.98 × 10^−6^; LAVmax_*i*_: *P* = 1.04 × 10^−3^).

**Figure 2.**
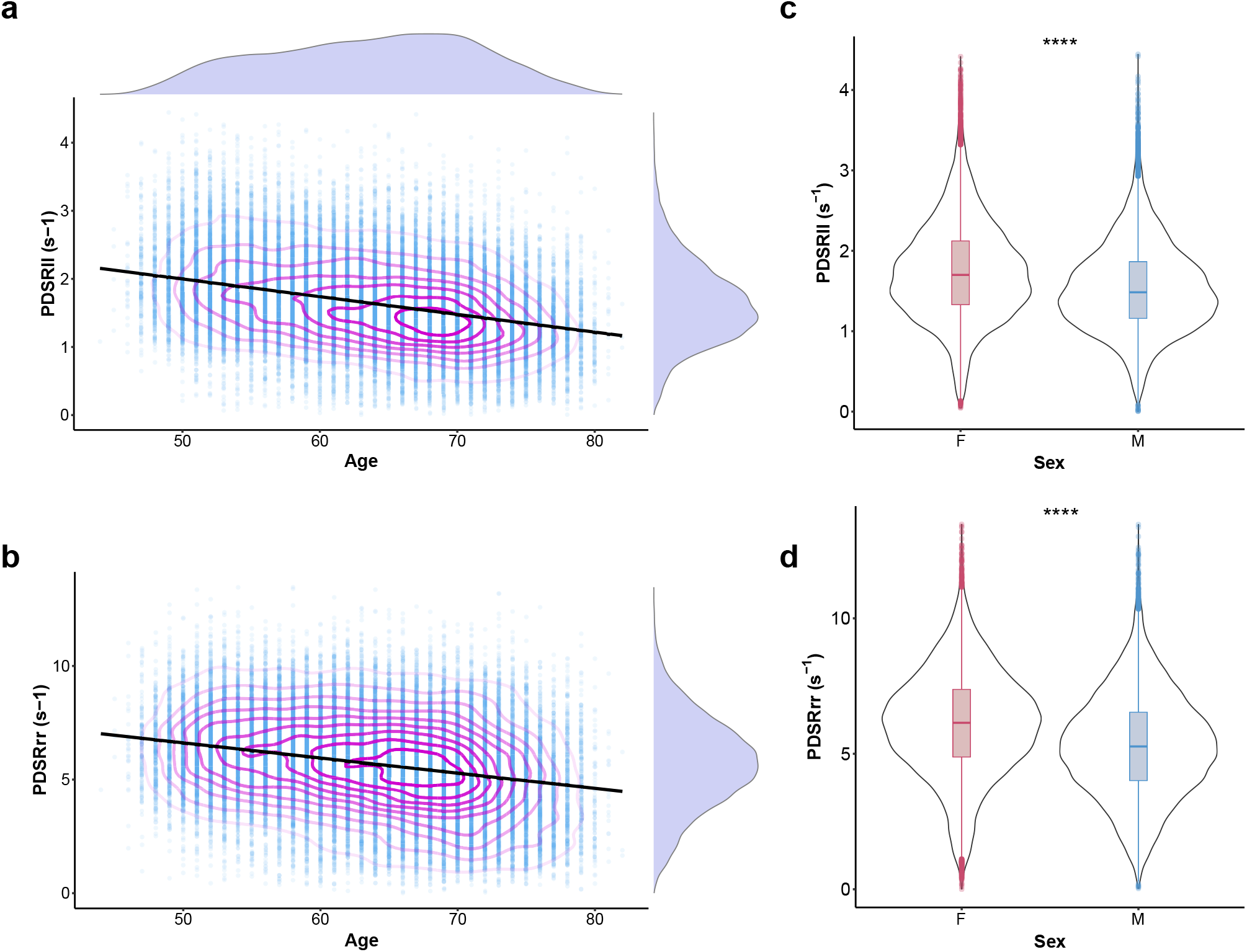
Population strain data. Scatterplots of a) longitudinal (*n* = 38, 923) and b) radial peak diastolic strain rates (PDSR_*ll*_ and PDSR_*rr*_ respectively) with age (*n* = 38, 700); with density contours, linear model fit and marginal density plots. Violin plots of c) longitudinal (*n* = 38, 923) and d) radial (*n* = 38, 700) peak diastolic strain rates with sex; \*\*\*\**P* < 0.0001, with boxplots showing the median, hinges showing the inter quartile range (IQR), and whiskers 1.5 x IQR.

**Figure 3.**
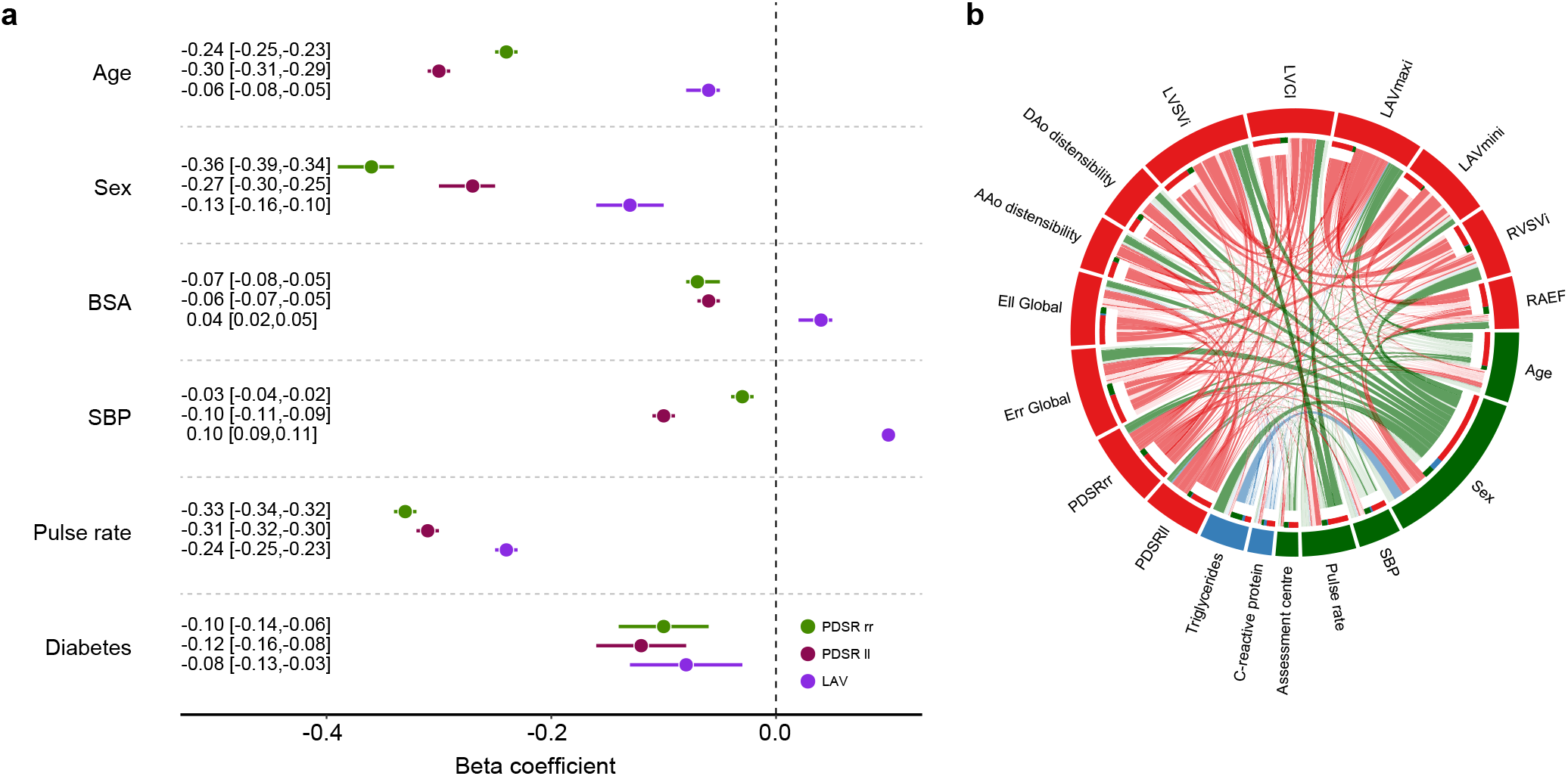
Regression analysis. a) Multiple linear regression analysis of left ventricular longitudinal (PDSR_*ll*_) and radial (PDSR_*rr*_) peak diastolic strain rates and indexed left atrial maximum volume (LAVmax_*i*_) with age, sex body surface area (BSA) systolic blood pressure (SBP), pulse rate and diabetes as predictors. All associations were significant after false discovery rate correction. b) Circular plot visualisation of the associations between the imaging (red - PDSR_*ll*_, PDSR_*rr*_, global systolic radial strain (E_*rr*_), global systolic longitudinal strain (E_*ll*_), ascending aortic (AAo) distensibility, descending aortic (DAo) distensibility, indexed left ventricular stroke volume (LVSV_*i*_), left ventricular cardiac index (LVCI), LAVmax_*i*_, indexed right ventricular stroke volume (RVSV_*i*_), and right atrial ejection fraction (RAEF)) and the non-imaging phenotypes (green for environmental; blue for biochemical). The strength of the connection between each pair is presented as a ribbon, whose size is proportional to their regression coefficient. All associations with a regression coefficient <0.3 are shown in faint colours (apart from the associations between PDSR_*ll*_, PDSR_*rr*_ and LAVmax_*i*_ and all other phenotypes). Standardised beta coefficients are shown with units in standard deviations for each variable.

To investigate the association between a comprehensive set of image-derived measures of atrial, ventricular and aortic function with a broader range of non-imaging phenotypes we used regression analysis with variable selection to fit a sparse model of predictors (Extended Data Fig 4). For this analysis, we excluded 7,936 subjects with incomplete data. The final model described the relationship between 12 imaging phenotypes, 5 non-imaging phenotypes and 2 serum biomarkers (Figure 3b) (see Supplementary Material for further information). Sex was associated with several systolic and diastolic parameters, including aortic function, as well as higher serum triglycerides in men. C-reactive protein (CRP), a circulating biomarker of inflammation, showed a positive relationship with serum triglycerides, but we found no circulating biomarkers independently associated with diastolic function. We found that reduced peak diastolic strain rates were associated with reduced LAVmax_*i*_. Left atrial function was related to indicators of right ventricular function emphasising their functional interdependence^15^.

### Genetic architecture of diastolic function traits

#### Genome-wide association analyses of diastolic function traits

The SNP-based heritability (i.e. the proportion of variance per trait explained by all considered SNPs) was 12% for PDSR_*ll*_, 13% for PDSR_*rr*_, and 21% for LAVmax_*i*_. The observed genetic correlation between the diastolic function traits was 0.22 (SE 0.07) between PDSR_*ll*_ and LAVmax_*i*_, 0.12 (SE 0.08) between PDSR_*rr*_ and LAVmax_*i*_, and 0.85 (SE 0.04) between PDSR_*ll*_ and PDSR_*rr*_.

Within the discovery set, we identified 5 independent loci (LAVmax_*i*_, 1; PDSR_*rr*_, 3; PDSR_*rr*_, 1) reaching genome-wide significance (*P* = 5·10^−8^; Supplementary Fig 3), which were also significant in the validation dataset also (*P* < 0.05*/*5). Considering the full dataset, the number of significant independent loci increased to 9 with 2 additional loci associating with PDSR_*rr*_, 1 additional with LAVmax_*i*_, and 1 additional with PDSR_*ll*_ (Figure 4).

**Figure 4.**
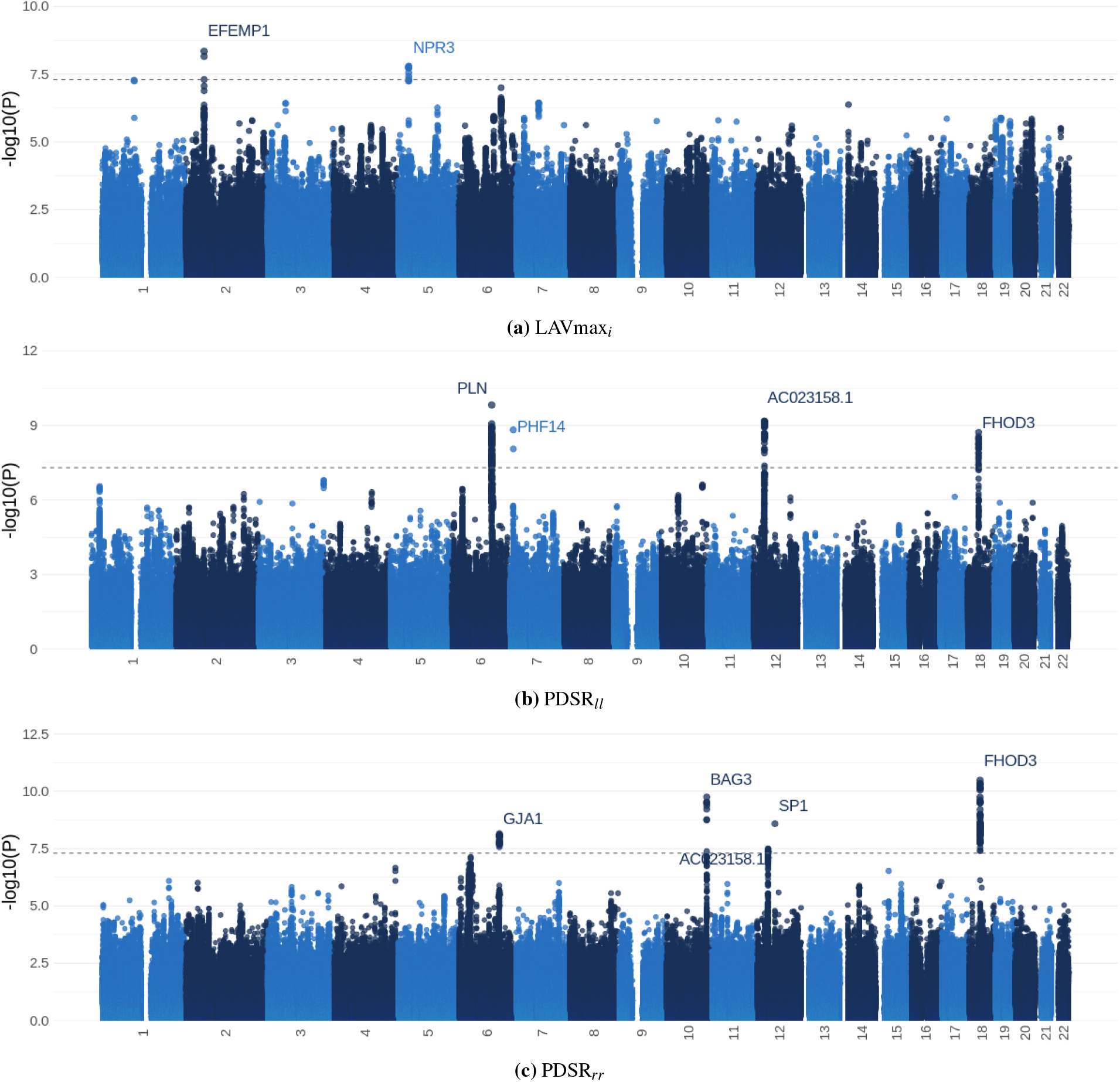
Manhattan plots of the GWAS results for the three diastolic function traits (full dataset). This figure shows the -log10(P-value) on the y-axis across all autosomal chromosomal positions (x-axis) from BOLT-LMM. The dotted line indicates genome-wide significance (*P* = 5 · 10^−8^, *N* = 34245). Significant loci are labeled by their likely causal gene, see Table 1.

#### Variant annotation

Summary information for the 9 loci identified using the full dataset is presented in Table 1 (further information can be found in Supplementary Material, Supplementary Fig 5 and Supplementary Table 1). The closest gene to each locus is depicted, with further variant to gene mapping presented as the “likely gene” given by evidence of a functional effect on a gene (details in Supplementary Material), additional heart-related phenotype associations, or a previously reported mechanism linking the gene to diastolic function. Through this approach we were able to highlight several structural genes associated with diastolic function that also have a known role in myocardial contractility (e.g. *TTN, PLN, GJA1*), and in the functional maintenance and stress-response of the cytoskeleton (eg *FHOD3, BAG3*)^16^. Moreover, we were also able to identify a novel link between the *NPR3* locus and left atrial volume. The signal co-localizes with a previously discovered association with blood pressure traits (systolic, diastolic and mean arterial blood pressure). The C-allele of the lead SNP (rs1173727) at this locus increases *NPR3* expression, and is associated with increased blood pressure and LAVmax_*i*_, and a nominally significant increase in risk of heart failure (Supplementary Material). The *NPR3* gene encodes the C-type natriuretic peptide receptor, which has a high tractability score (Table 1), making it an attractive therapeutic target.

**Table 1.**
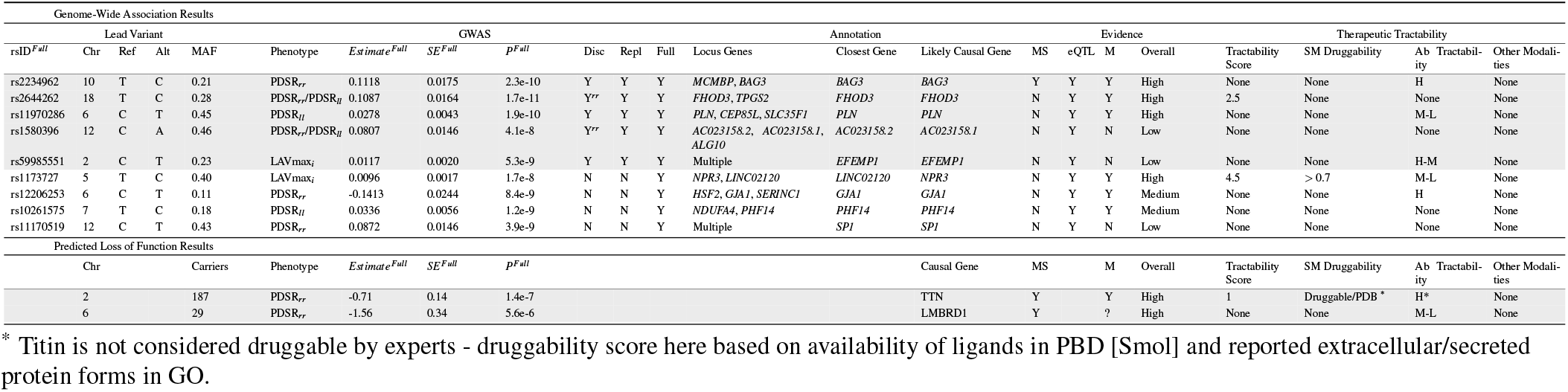
Summary information on the lead variants identified from each GWAS analysis and the significant genes from the Loss of Function analysis. For each significant locus across the 3 diastolic phenotypes, variant information, GWAS summary statistics, variant to gene annotation, and potential therapeutic tractability of the likely causal gene is provided. The evidence column is split by: MS - missense variant; eQTL - colocalisation between the GWAS signal and an eQTL for the gene in a plausible tissue type (see Supplementary Material); M - plausible mechanistic link between the gene and the measured heart phenotypes i.e. the gene function suggests a link to diastolic function; Overall - the confidence of variant to gene mapping given all the available evidence. Therapeutic tractability is split into 4 categories: tractability score - the Ensembl tractability score; SM druggability - a categorical assessment of small molecule tractability (low, medium, high); Ab tractability - a categorical assessment of antibody tractability (low, medium, high); Other modalities - enzyme, protein, oligonucleotide, etc.^19^. Loci highlighted in grey are those that reached genome-wide significance in the discovery, validation, and full datasets, loci in white reach suggestive significance in the discovery dataset and genome-wide significance in the full dataset. Further information is provided in the Supplementary Material.

NPR3 accounts for almost 95% of the natriuretic peptide receptor (NP) population^17^. It binds all 3 members of the natriuretic peptide pathways, natriuretic peptides A, B and C (ANP, BNP and CNP) and has been ascribed both a clearance role for these peptides, regulating their circulating levels, and a signalling role for CNP^18^. To further elucidate the biological relationship between the lead *NPR3* SNP and heart failure, we first sought to study in more detail individuals with predicted loss-of-function variants in *NPR3* or its ligands but, within the group with whole-exome sequencing data, the numbers of such individuals were limited (*NPR3* n=5, *nNPPA* n=2, *nNPPB* n=2, *nNPPC* n=0).

We next examined the relationship between common variants in *NPR3* and genes encoding other proteins in the natriuretic peptide pathway with traits linked to the lead SNP, rs1173727. Full results of this systematic assessment are shown in Supplementary Fig 6, and an abridged version is provided in Extended Data Fig 6. We identified additional genetic variants in *NPR3* with robust effects on other traits influenced by the LAVmax_*i*_ associated variant rs1173727. Hierarchical clustering of the variant-trait effects showed that the observed effects for the *NPR3* variant overlap with both the *NPPA* & *NPPB* genes and the effects of genetic variation in *NPPC* or *NPR2*. In brief, we were unable to distinguish between the two proposed roles for *NPR3* in the context of this study.

### Potential causes and consequences of diastolic function

#### Creation of polygenic risk scores (PRS and PheWAS)

PRSs for each diastolic function trait consisted of 20 SNPs for PDSR_*rr*_, 15 SNPs for PDSR_*ll*_, and 8 for LAVmax_*i*_. The PRS explained 1.5 % of the variability of PDSR_*rr*_, 1.1 % of PDSR_*ll*_ and 0.2 % of LAVmax_*i*_. There was good agreement between the distribution of the PRS in the UK Biobank participants with and without CMR indicating no systematic bias in genetic architecture (Supplementary Fig 8). The Pearson correlation coefficient for the PRS for PDSR_*ll*_ and PDSR_*rr*_ was 0.35 whereas the correlation coefficient between LAVmax_*i*_ and PDSR_*ll*_ or PDSR_*rr*_, respectively, was much lower (< 0.01). PheWAS was undertaken and we considered traits that have been previously associated with cardiac phenotypes in the literature, but in addition included an unbiased selection of phenotypes for exploration. In total, we considered 71 quantitative phenotypes and 63 (binary) disease endpoints (Supplementary Data 1). Out of these, 31 phenotypes were significantly associated (Padj<0.05) with at least one of the diastolic function PRSs after leave-one-out cross validation (Figure 5). Some of the identified PheWAS associations are consistent with the phenotype correlation analysis (e.g. pulse rate and blood pressure). We also confirmed associations between diastolic function and previously reported biomarkers of heart failure (e.g. SHBG^20^ and IGF-1^21^). Furthermore, we identified an association of PDSR_*rr*_ to heart failure, cardiomyopathy and dilated cardiomyopathy, implicating diastolic function in cardiovascular endpoints.

**Figure 5.**
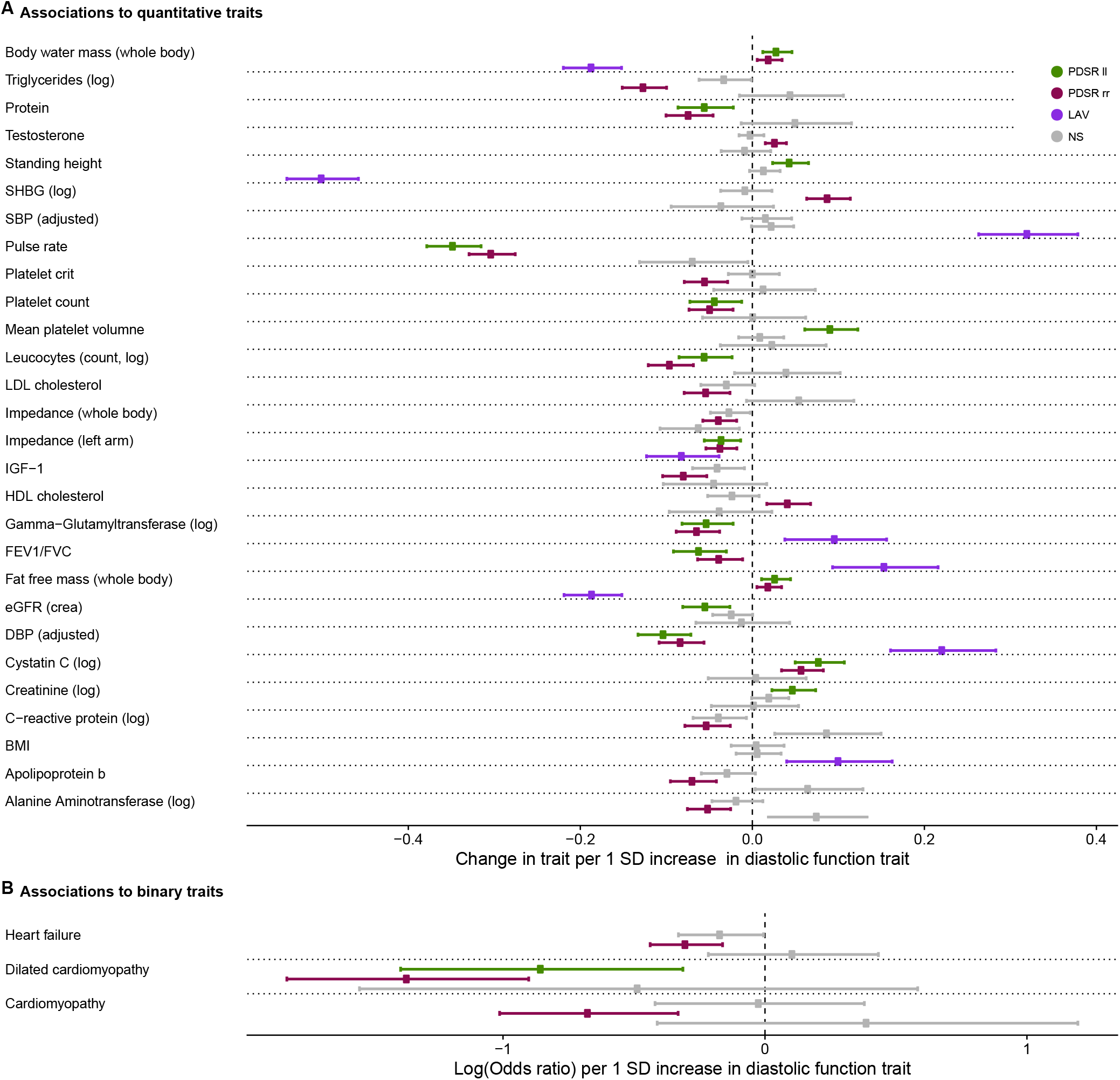
Significant associations of the PRSs for diastolic function traits with UK Biobank phenotypes. A) Quantitative traits that significantly associated with the PRSs of diastolic function. B) Binary traits that significantly associated with the PRSs of diastolic function. Numerical results, including P-values and 95 % confidence intervals are shown in Supplementary Table 4, Supplementary Table 5, and Supplementary Table 6. One unit change in the PRS represents a change of 1 standard deviation in the respective diastolic function trait. All dependent variables (traits) were standardized representing the change in dependent variable standard deviations for a 1-standard-deviation change in the respective measurement. Associations not significant after multiple testing correction (conducted per PRS) are displayed as grey bars. SBP: systolic blood pressure, MAP: mean arterial pressure, DBP: diastolic blood pressure, Log: natural logarithm, NS: non-significant. *N* = 449263.

#### Mendelian randomization

We used Mendelian Randomization (MR) to identify causal bi-directional relationships between diastolic function and cardiovascular outcomes (heart failure, diabetes, pulse rate, and diastolic blood pressure) which we chose based on clinical plausibility and the findings of the phenotype correlation analysis. As we hypothesize LAVmax_*i*_ to be downstream of PDSR_*rr*_ and PDSR_*ll*_, reflecting that atrial volume is consequent on ventricular filling^22^, we focussed on PDSR_*rr*_ and PDSR_*ll*_ while results for *LAV max*_*i*_ are reported for completeness in Supplementary Table 9 and in Supplementary Fig 10. We identified a potential causal, bi-directional association between pulse rate and both PDSR_*rr*_ and PDSR_*ll*_ (Figure 6, Supplementary Table 7, Supplementary Table 8), supporting findings from pre-clinical models suggesting that selective heart rate reduction can affect diastolic function^24^. We also identified a likely causal relationship between lower PDSR_*rr*_ (stiffer ventricle) and increased risk of heart failure Supplementary Fig 9, which was further corroborated using GWAS summary results^25^ from the HERMES consortium (Supplementary Table 10), which is a GWAS meta-analysis from 47309 heart failure cases and 930014 controls. The magnitude of the effect observed in the MR analysis is consistent with the observational epidemiological estimate, derived from correlating PDSR_*rr*_ with incident heart failure (Extended Data Fig 5). We found no causal relationship between longitudinal PDSR_*ll*_ and heart failure, and neither was one observed in our epidemiological analysis (Extended Data Fig 5). Diastolic dysfunction is frequently present in diabetic patients^26^, but although we observed a difference in PDSR_*rr*_ between diabetics and non-diabetics (Figure 3), we could not confirm a causal relationship between diabetes as an exposure with either PDSR_*ll*_ or PDSR_*rr*_ as outcomes (Supplementary Table 7, Supplementary Table 8).

**Figure 6.**
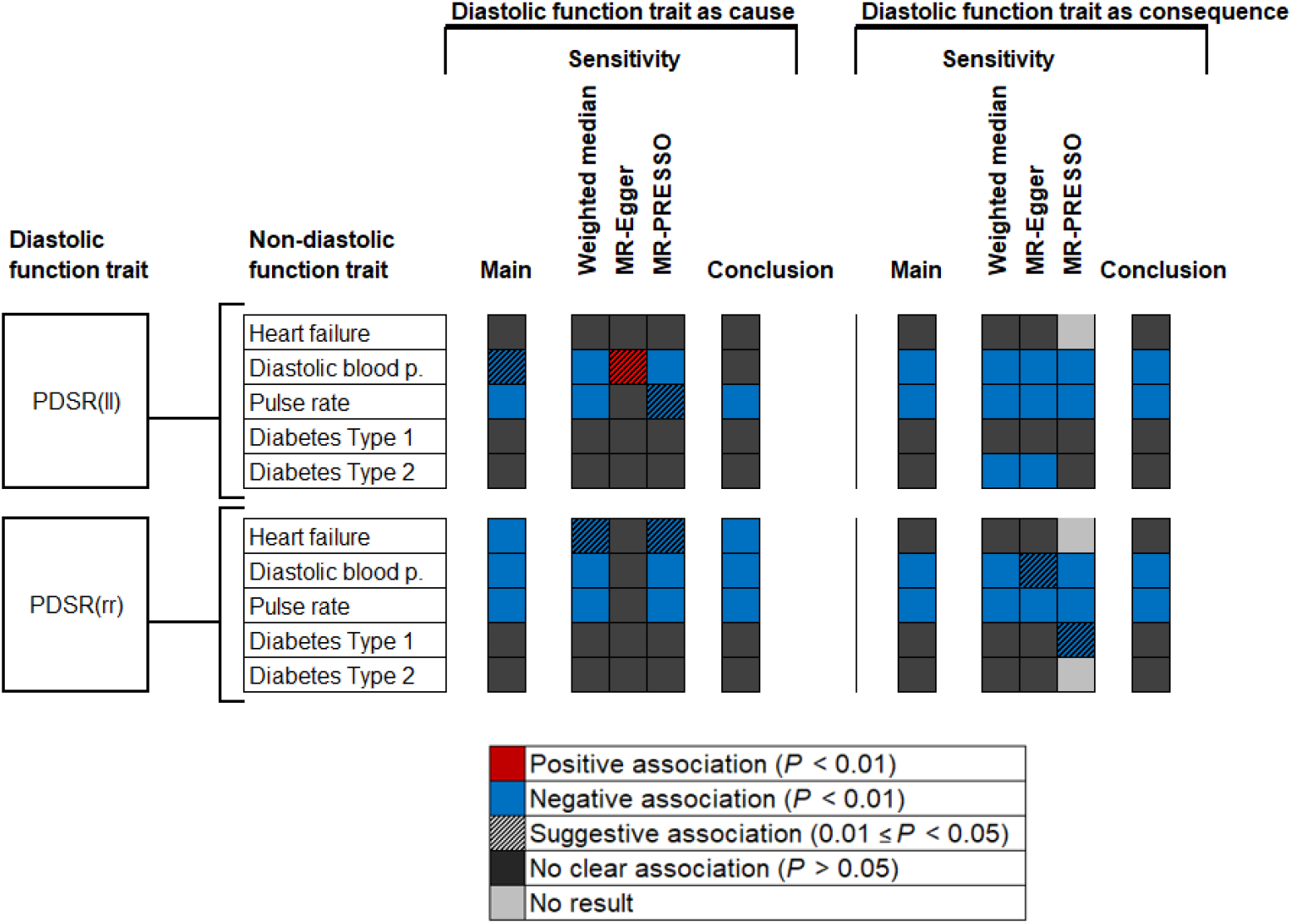
Summary of results from MR analyses of traits describing diastolic strain and multiple cardiac outcomes and phenotypes. The style of this figure is adapted from van Oort et al.^23^.“No result”, no outlier was detected with MR-PRESSO and therefore the MR-PRESSO estimate was identical to the IVW estimate. Detailed results are provided in Supplementary Table 7 and Supplementary Table 8

## Discussion

Diastole is a complex series of molecular, biophysical and electro-mechanical processes that initiate contractile deactivation and promote efficient ventricular filling. Impairment of these coordinated mechanisms may lead to diastolic dysfunction which is associated with the presence of multiple cardiovascular risk factors leading to reduced quality of life and higher mortality^27,28^. Here, we used deep learning cardiac motion analysis to perform the first reported GWAS of diastolic function traits with the aim of determining tractable causative mechanisms. We found that diastolic function was a heritable trait with associations in loci related to myofilament mechanics, protein synthesis during mechanical stress and regulation of cardiac contractility. Furthermore, we find a role for endothelium-derived signalling in diastolic function that is a potential therapeutic target^29^. Lastly, through Mendelian randomisation we observe a causal relationship between genetically-determined diastolic function and heart failure outcomes.

A decline in diastolic function is a feature of the ageing heart and we found that age was a strong independent predictor of diastolic function, with a greater decrease present in males. Outcome studies have suggested that this is a prognostically-benign feature of healthy ageing that is not related to adverse effects of cardiac senescence^4,30,31^. Changes in titin protein phosphorylation, myocardial redox state and impairment of nitric oxide signalling have been proposed as potential mechanisms^32^, and clinical studies indicate that age-related myocardial fibrosis, cardiomyocyte hypertrophy, and reduced microvascular density, may be a consequence rather than an initiating cause of diastolic dysfunction^33^.

We found that diabetes was associated with impaired diastolic function and this relationship was independent of age, BSA, and systolic blood pressure. Increased myocardial stiffness is recognised as one of the earliest, and potentially reversible, manifestations of myocardial dysfunction in diabetes^34^. Several underlying mechanisms related to insulin resistance have been proposed that include altered cardiac energetics and accumulation of advanced glycation end products that promote ventricular stiffness^35^. However, our findings did not conclusively point to a causal relationship between diabetes and diastolic function using Mendelian randomisation. We observed a unidirectional causal relationship of PDSR_*rr*_ on heart failure risk, as well as associations with cardiovascular endpoints and circulating biomarkers of heart failure through PheWAS. Longitudinal cohort studies have suggested that persistence or progression of diastolic dysfunction is a risk factor for subsequent heart failure^36^, and our findings suggest that ventricular stiffness is a substrate for the evolution of mixed aetiology heart failure. In contrast, our findings show that variation in left atrial volume (*LAV max*_*i*_) is likely a consequence of myocardial diastolic function which is not itself causally related to heart failure risk.

Our study provides insights into the biological basis of diastolic function with potential implications for therapy development.

We identified common variants within genes implicated in cardiomyopathies (e.g. *BAG3, FHOD3, PLN*), suggesting sarcomere homeostasis during mechanical stress may affect diastolic function in both health and disease^37^. A further example is the identification of the gene of a key regulator of cardiac diastolic function, phospholamban (*PLN*), which modulates sarcoplasmic reticulum calcium-ATPase activity^38^. We also identified a potential therapeutic target through the association of DNA variants at the locus of *NPR3* influencing diastolic function and risk of heart failure. Previous studies have highlighted its role in blood pressure control^39^. NPR3 has been described as a clearance receptor for ANP, BNP and CNP, while more recently as a functional signalling receptor for CNP has become apparent^29^. A recent mechanistic study in animal models^29^ identified a role for this receptor in mediating the cardioprotective effects of cardiomyocyte and fibroblast-released CNP. While we cannot exclude a signalling function for NPR3, the direction of effect in our genetic analysis is such that reduced *NPR3* gene expression is associated with reduced LAVmax_*i*_, blood pressure and heart failure risk. This is consistent with its role as a clearance receptor, where reduced expression would enhance exposure to the beneficial effects of ANP and BNP and outweigh loss of CNP activity. Inhibition of natriuretic peptide degradation via inhibition of neutral endopeptidase is already an approved licensed treatment for heart failure, validating the pathway as a therapeutic target. Our observations support pharmacological inhibition of NPR3 as an additional approach to managing this condition.

This analysis has some limitations. UK Biobank is a large-cross sectional study that is subject to selection bias and latent population stratification^40^, however risk factor associations appear to be broadly generalisable^41^. The population is predominantly European and further work is required to explore diastolic traits and outcomes in people of diverse ancestries. Echocardiography has been the cornerstone of assessing diastolic function by characterising features of ventricular relaxation, stiffness and recoil^2^. However, feature-tracking CMR has excellent agreement with speckle-tracking echocardiography^42^ and invasive measures of diastolic function^43^.

In conclusion, we found that diastolic function is a heritable trait that is causally upstream of heart failure. Associated common variants are related to genes that maintain functional homeostasis under biomechanical stress. We also identify a gene encoding an atrial natriuretic peptide receptor as a novel therapeutic target for modulating aspects of diastolic function.

## Methods

All analyses in this study can be found here https://github.com/ImperialCollegeLondon/diastolic_genetics/, and were conducted with R version > 3.6.0.

### Participants

For UK Biobank, approximately 500,000 community-dwelling participants aged 40–69 years were recruited across the United Kingdom between 2006 and 2010^44^. All subjects provided written informed consent for participation in the study, which was also approved by the National Research Ethics Service (11/NW/0382). Our study was conducted under terms of access approval number 28807 and 40616. A range of available data were included in this study comprising genotyping arrays and whole exome sequencing, cardiac imaging, health-related diagnoses, and biological samples.

There are 488,252 genotyped participants of which 200,640 have whole exome sequencing. We partitioned 39,559 participants with both CMR imaging and genotyping array data into two tranches by date of release from UK Biobank providing a discovery dataset of 26,893 participants and a validation dataset of 12,666 participants.

### Imaging protocol

A standardised CMR protocol was followed to assess cardiac structure and function using two-dimensional retrospectively-gated cine imaging on a 1.5T magnet (Siemens Healthineers, Erlangen, Germany). A contiguous stack of images in the left ventricular short-axis plane from base to apex was acquired, with long axis cine imaging in the two and four chamber views. Each cine sequence had 50 cardiac phases with an acquired temporal resolution of 31 ms^10^. Transverse cine imaging was also performed in the ascending and descending thoracic aorta. Images were curated on open-source databases^45,46^. All imaging phenotypes used for the analysis underwent quality control assessment^11^. Participants also underwent a resting 12 lead electrocardiogram which was automatically analysed using proprietary software (CardioSoft, GE Healthcare).

### Cardiac image analysis

Segmentation of the short-axis and long-axis cine images in UK Biobank was made using fully convolutional networks, a type of deep learning neural network, which predict a pixel-wise image segmentation by applying a number of convolutional filters onto each input image for feature extraction and classification^12^. The accuracy of image segmentation on the UK Biobank dataset is equivalent to expert human readers^47^. End-diastolic volume, end-systolic volume, stroke volume, and ejection fraction were determined for both ventricles. Left ventricular myocardial mass was calculated from the myocardial volume assuming a density of 1.05 g.ml ^−1^. Left atrial volume was calculated from the segmented images using the biplane area–length formula 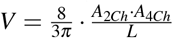, where *A*_2*Ch*_ and *A*_4*Ch*_ indicate the atrial area on the two and four-chamber cines respectively, and *L* indicates the longitudinal diameter averaged across two views. Measurements were indexed to body surface area (BSA) according to the Du Bois formula: 0.20247 (*Weight*^0.425^) (*Height*^0.725^), with weight in kg and height in m. The heart was divided into 16 standardised anatomical segments, excluding the true apex, according to American Heart Association nomenclature^48^.

The aorta was segmented on the cine images using a spatio-termporal neural network^49^. The maximum and minimum cross-sectional areas were derived from the segmentation and distensibility calculated using estimates of central blood pressure obtained using peripheral pulse-wave analysis (Vicorder, Wuerzburg, Germany)^11^.

Motion tracking was performed on the cine images using non-rigid image registration between successive frames (in GitHub repository ukbb_cardiac)^50,51^. To reduce the accumulation of registration errors motion tracking was performed in both forward and backward directions from the end-diastolic frame and an average displacement field calculated^11^. This motion field was then used to warp the segmentation contours from end-diastole onto successive adjacent frames. Circumferential (*E*_cc_) and radial (*E*_rr_) strains were calculated on the short axis cines by the change in length of respective line segments (Figure 1A) as 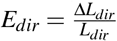, where *dir* represents the direction, *L*_*dir*_ the length of a line segment along this direction and Δ*L*_*dir*_ its change over time. Motion tracking was also performed on the long-axis four-chamber cines to derive longitudinal (*E*_ll_) strain. Peak strain for each segment and global peak strain were then calculated (Figure 1B). Strain was measured from slices acquired at basal, mid-ventricular, and apical levels. For comparison between each component absolute strain values are reported^52,53^. Strain rate was estimated as the first derivative of strain and peak early diastolic strain rate in radial (PDSR_rr_) and longitudinal (PDSR_ll_) directions was detected using an algorithm to identify local maxima (in GitHub repository peak_detection) (Figure 1C).

### Non-imaging phenotypes

In total we consider 110 non-imaging cardiovascular-related phenotypes in UK Biobank participants for the phenotype regression analysis and the genetic analysis. These phenotypes contain information acquired by touch screen questionnaire, interview, biophysical measurement, hospital episode statistics, primary care data and biochemical analysis of venous blood. Details of how each phenotype was acquired are available on the UK Biobank Showcase (http://biobank.ctsu.ox.ac.uk/crystal/). It should be noted that the biochemical markers used here were acquired at the initial assessment visit that preceded imaging assessment. Also, note that not all phenotypes were used in both the phenotype and the genetic analysis (e.g., due to lack of available data at the imaging visit). We refer to the Supplementary Material both for details on the definition of the considered phenotypes and for information on the inclusion of specific phenotypes for each analysis.

### Statistical significance testing and multiplicity control

We consider in all phenotype analysis a *P*-value < 0.05 as significant. Where not stated otherwise, we control the false discovery rate with the Benjamini-Hochberg adjustment^54^. Significance thresholds and decision criteria for GWAS significant loci and causality assessment (Mendelian Randomization) are described in the respective sections and/or in the Supplementary Material.

### Phenotype Association Analysis

Continuous variables are expressed as mean ± standard deviation (SD). Differences in continuous variables between groups were performed using Student’s t-test. Univariable and multiple linear regression analysis was used to explore the phenotype relationship between each diastolic parameter and cardiovascular risk factors. To identify relationships between diastolic function and a broader range of imaging and non-imaging phenotypes, including circulating biomarkers, we used the least absolute shrinkage and selection operator (LASSO)^55^ with stability selection, to optimise the model coefficients. We then ran regression diagnostics on the model with the selected variables, to exclude a possible collinearity inappropriately influencing our model (see Supplementary Material for details on the phenotype analysis and LASSO analysis procedure).

### Genotyping and sample QC

Genotyping of UK Biobank participants has been described elsewhere in detail^56^. Briefly, UK Biobank genotyping for 488,252 subjects was performed on the UK BiLEVE or UK Biobank Axiom arrays. Imputation was based on the HaplotypeReference Consortium panel and the UK10K+1000 Genomes panel^56^. In this study, UK Biobank Imputation V3 (in GRCh37 coordinates) were used. Whole exome sequencing (WES) was performed on data released in 2020 collected from 200,640 UK Biobank participants^57^. The sequencing methods and variant calling procedures have been described in detail^58^. In the present study, genotypes in their released PLINK-format files are utilized, and samples were restricted to the European population. Quality control of the genetic data was performed as recommended by UK Biobank (see Supplementary Material for details on the procedure and number of excluded samples).

### GWAS analysis

For the genetic analysis, there were 34,242 participants of European ancestry (see Supplementary Material for criteria) providing a discovery dataset of 23,321 participants and a validation set of 10,924 participants. GWAS analyses for the three diastolic function traits and additional quantitative traits of interest (as described for the causality assessment) were performed with BOLT-LMM (version 2.3.2) which accounts for ancestral heterogeneity, unknown population structure, and sample relatedness^59,60^. GWAS analyses were adjusted for imaging traits for the first ten genetic principal components, sex, age at time of MRI, the genotyping array and the MRI assessment center and for non-imaging quantitative traits for the first ten principal components, sex, age at measurement of the trait and the genotyping array. GWAS analyses for clinical endpoints of interest (binary endpoints) were conducted with PLINK2^61^ and adjusted for the first ten principal components, sex, age at baseline and the genotyping array. Post-GWAS filtering removed any SNPs with a Hardy-Weinberg equilibrium p-value < 0.05 and *MAF* < 0.005.

### Assessment of shared genetic architecture

For the assessment of shared genetic architecture between diastolic function traits, LD score regression (LDSC (LD SCore) v1.0.1, PMID 25642630) was used to obtain a genetic correlation score between each pair of traits.

### Variant annotations

Lead variants for each locus were assigned causal genes, where possible, using a combination of variant annotations and additional functional genomic data sources (colocalisation). Each lead variant was systematically tested for any evidence of functional consequence using VEP. In addition, QTL evidence was extensively searched using Open Targets Genetics^62^. Where eQTL data was available for the locus, the full summary statistics were downloaded to assess colocalisation (see Suppplementary Material).

Variant Effect Predictor (VEP)^63^ and Loss-of-Function Transcript Effect Estimator (LOFTEE)^64^ plugin were applied on all genomic variants of WES data. In the present study, we considered the genomic variants predicted by LOFTEE with high-confidence label “HC”, non-dubious (no “LoF flag” such as variants that located in poorly conserved exons, or splice variants that affect NAGNAG sites or non-canonical splice regions), and minor allele frequency < 0.05, as a Loss-of-function (LoF) mutation.

### LoF association analysis

A Loss-of-Function (LoF) carrier indicator was created for each WES sample and each of the human protein-coding genes based on the collapsed information of LoF annotations. A subject was considered as an LoF carrier of the gene if there was at least one LoF mutation (based on methods in the variant annotation section), and a non-carrier if there was none. We then conducted the association test between LoF carrier indicator and the three diastolic function imaging phenotypes. Linear regression was performed with the adjustment of sex, age at time of MRI, and the top ten genetic principal components. The association results were further filtered as those with at least two carriers and the endpoint available. The association was considered significant after multiple testing correction at *α* = 0.05 (FDR, calculated for three diastolic function traits). We identified 18,660 participants with both whole exome sequencing data and CMR imaging data.

### Polygenic risk scores (PRS)

Candidate variants for PRS for the three diastolic function traits (LAVmax_*i*_, PDSR_*ll*_, PDSR_*rr*_) were obtained based on the respective GWAS (full imaging cohort) results by performing clumping (PLINK1.9,^65^) using an LD threshold of *R*^2^ = 0.1 (in a window of 1000kb) and considering all SNPs with *P* < 10^−6^. Candidate variants were included in multivariate linear modelling evaluated on the European subset of the full imaging cohort with the first ten genetic principal components, age at MRI, sex, genotyping array and the MRI center as additional covariates and the respective diastolic function trait as dependent variables. The diastolic function traits were scaled to standard deviation (sd) 1 prior to the model estimation - therefore, a unit change in the PRS score represents a change of 1 sd unit in the respective diastolic function trait. PRS estimates per individual were then calculated by multiplying the observed genotype with the estimated beta from the multivariate linear model for each SNP and summing these values up. Missing genotypes were imputed using a mean imputation. The variance explained for the PRS is measured by *R*^2^, estimated in a linear regression with the PRS as only variable and the respective diastolic function trait as endpoint.

Next, we conducted a PheWAS using the obtained PRS (see above and Supplementary Material for a full definition of included phenotypes in the PheWAS). Evaluation of the PRS were performed in the European non-imaging cohort, i.e. an independent set of subjects compared to the PRS construction set. Only results are shown that are significant after multiple testing correction at *α* = 0.05 (FDR, calculated per diastolic function trait) and, as a sensitivity analysis, for which all leave-one-SNP out cross validations analysis lead to a significant result at *α* = 0.05 after multiple testing correction (FDR) for the number of considered phenotypes. The latter condition is supposed to exclude spurious results which are only driven by one single variant. Leave-one-SNP out cross-validation is performed by excluding one SNP from the list of candidate variants, then re-estimating the PRS and performing the PheWAS as described above. For the leave-one-SNP out cross-validation, FDR adjustment is performed per combination of diastolic trait and phenotype, considering the number of included SNPs.

### Mendelian randomization

For exploring the causes and consequences of diastolic function parameters, we used a bidirectional Mendelian randomization (MR) approach, i.e. two MR analysis are performed: first, an MR analysis using the first chosen trait as exposure is conduced and secondly a MR analysis using the selected second trait is run. By considering both results, evidence can be gathered for a one-directional causal relationship, a bi-directional causal relationship or no causal relationship at all^66^. We performed this analysis taking into account one diastolic and one non-diastolic function trait and for that, we selected non-diastolic function traits of interest by taking into account the results from the observational correlation analysis and clinical expertise. This approach lead to the consideration of 5 traits (Heart failure, Diabetes - considering Type I and Type II separately-, pulse rate, diastolic blood pressure) as potential causes or consequences of changes in diastolic function parameters.

We established a workflow for the MR analysis which briefly described in this section. Full details are provided in the Supplementary Material. Genetic instrumental variables were selected from the UK Biobank GWAS results generated -as described above- via clumping with PLINK1.9 as described for the PRS approach. The candidate SNP set prior to clumping was restricted to the intersection between the SNP sets of the pair of GWAS results (hypothesised causal trait GWAS and hypothesised consequence trait GWAS).

All MR analysis are based on the point estimates and standard deviations obtained from the respective GWAS. We follow a similar approach to van Oort et al.^23^ by using inverse-variance weighted method (IVW) as the main analysis and applying several other MR methods for ensuring robustness of the obtained results as sensitivity analyses. We used weighted median-based methods^67^, MR-PRESSO^68^ and MR-Egger^69^. Consistent effect estimates across the different methods improves our confidence in a truly causal effect. We consider an association as “potential causal” if the main analysis indicates a causal relationship (*P* < 0.01), at least two of the sensitivity analyses indicate at least a suggestive causal relationship (*P* < 0.05) and none of the sensitivity analyses indicate associations with inconsistent effect directionality, i.e. none of the methods showed a suggestive association with conflicting directionality (*P* < 0.05). No explicit multiplicity adjustment is performed for MR experiments. For “potential causal” associations, we next conducted a supplementary sensitivity analysis using published GWAS results as described in the Supplementary Material - if published GWAS data was available.

All analysis, which involved diastolic and non-diastolic function traits, were conducted in a two-sample approach, i.e., the diastolic function trait GWAS was calculated in the full imaging cohort and the non-diastolic function trait GWAS was calculated in the non-imaging cohort.

For comparison of the effect estimates from the MR-analysis to the observed correlation of diastolic function measurement and disease status, we restricted the analysis population to subjects which were disease-free at the CMR visit. We then fitted a logistic regression model by coding subjects who experienced a first event of the selected disease during follow-up time as 1 and event-free subjects during follow-up as 0. As covariates, we included age at CMR visit, gender, diabetes status, diastolic blood pressure and BMI. Note that this analysis was only performed for relationships judged as potential causal and that involves a disease endpoint (and not a quantitative measurement like pulse rate).

### *NPR3* Pathway Analysis

In order to increase our understanding of the association of NPR3 with LAVmax_*i*_ and to further characterize the role of natriuretic peptides, we looked for additional genetic associations within genes of the natriuretic peptide pathway (so in addition to *NPR3* - *NPR1, NPR2, NPPA, NPPB*, and *NPPC*). We conducted GWAS using BOLT-LMM for all imaging traits listed in Extended Data Table 1 as described above, as well as any non-imaging traits associated with rs1173727 (the lead variant for *NPR3*) across the 4 loci (*NPPA* and *NPPB* share the same locus). The GWAS summary statistics were filtered to a 1MB window around each gene (for *NPPA/B*, the gene used for centering was *NPPA*). Across these summary statistics, we performed clumping with a p-value threshold of 10^−5^ and *R*^2^ < 0.1.

For the identified tag SNPs and associated variants in LD from the clumping analysis, we then tested which of these variants we could confidently link to the natriuretic gene in the locus. If any variant was classified as missense, we selected that variant directly. For eQTL variants, we used colocalisation analysis to link these SNPS to the natriuretic genes in each locus. Relevant eQTL and pQTL data was used (eQTL summary statistics were taken from eQTL Catalog^70^ and pQTL data from Sun et al.^71^) and SNPs with only a clear association with the gene of interest and traits of interest were kept (i.e. *p* < 1^−4^ for association with gene or protein expression, *P* < 10^−5^ for association with the trait, and *H*_12_ > 0.5 was used as a threshold for the co-localization analysis).

Hierarchical clustering was then performed on the —log(*P*) × *β* values with the *β* s aligned to have a negative sign on the diastolic blood pressure. Extended Data Fig 6 shows all SNPs and traits with a genome-wide significant association. The SNPs and traits with suggestive associations (*P* < 10^−5^) are shown in the Supplementary Material (Supplementary Fig 6).

## Data Availability

All raw and derived data in this study is available from UK Biobank (http://www.ukbiobank.ac.uk/). GWAS summary level data are publicly available through the GWAS catalogue.

https://github.com/ImperialCollegeLondon/diastolic_genetics

## Acknowledgments

The study was supported by Bayer AG; Medical Research Council (MC-A658-5QEB0); National Institute for Health Research (NIHR) Imperial College Biomedical Research Centre; British Heart Foundation (NH/17/1/32725, RG/19/6/34387, RE/18/4/34215); Wellcome Trust (107469/Z/15/Z); Academy of Medical Sciences (SGL015/1006); Mason Medical Research Trust grant; and the Engineering and Physical Sciences Research Council (EP/P001009/1). H.V.M. also receives funding from the Simons Center for Quantitative Biology at Cold Spring Harbor Laboratory.

## Code Availability

The analysis code is freely available on GitHub (DOI: 10.5281/zenodo.4767044).

## Data Availability

All raw and derived data in this study is available from UK Biobank (http://www.ukbiobank.ac.uk/). GWAS summary level data are publicly available through the GWAS catalogue (accession number GCP000170).

## Author contributions

M.T. and J.M. performed the formal analyses and co-wrote the paper; W.B, N.S., A.de M. and D.R. performed the image analysis; K.A.McG., J.S.W., H.V.M., L.Z., F.S. and C.B. performed or interpreted the genetic analyses; A.MacN. performed the GWAS; M.R.W. interpreted the pharmacology findings; D.F.F. and D.P.O’R conceived the study, managed the project and revised the manuscript. All authors reviewed the final manuscript.

## Extended Data Figures

**Extended Data Figure 1.**
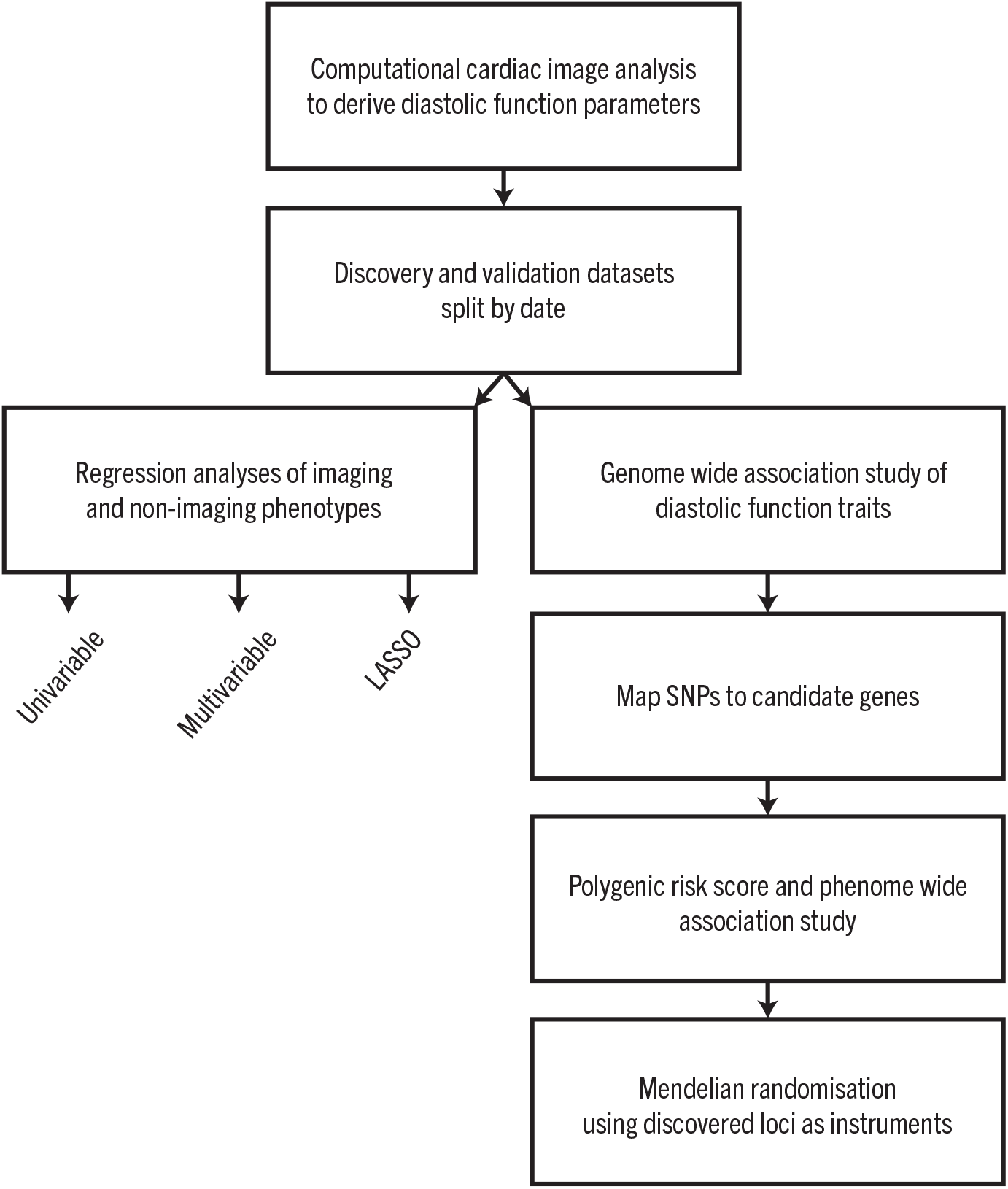
A summary of the main steps in our analysis of the genetic and environmental determinants of diastolic heart function.

**Extended Data Figure 2.**
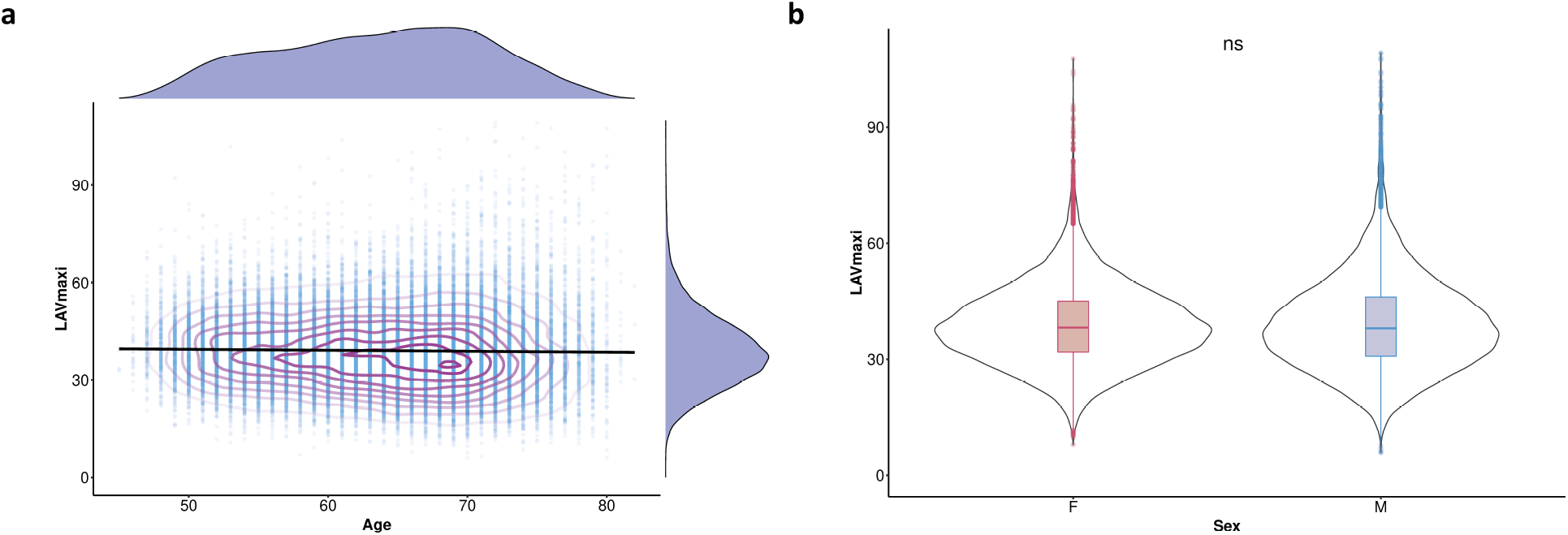
a) Scatterplots of left atrial maximum volume indexed to body surface area (LAVmax_*i*_) against age with density contours, linear model fit and marginal density plots. b) Violin plots of LAVmax_*i*_ by sex with boxplots showing median, interquartile range (IQR) and 1.5 x IQR (n=38,046)

**Extended Data Figure 3.**
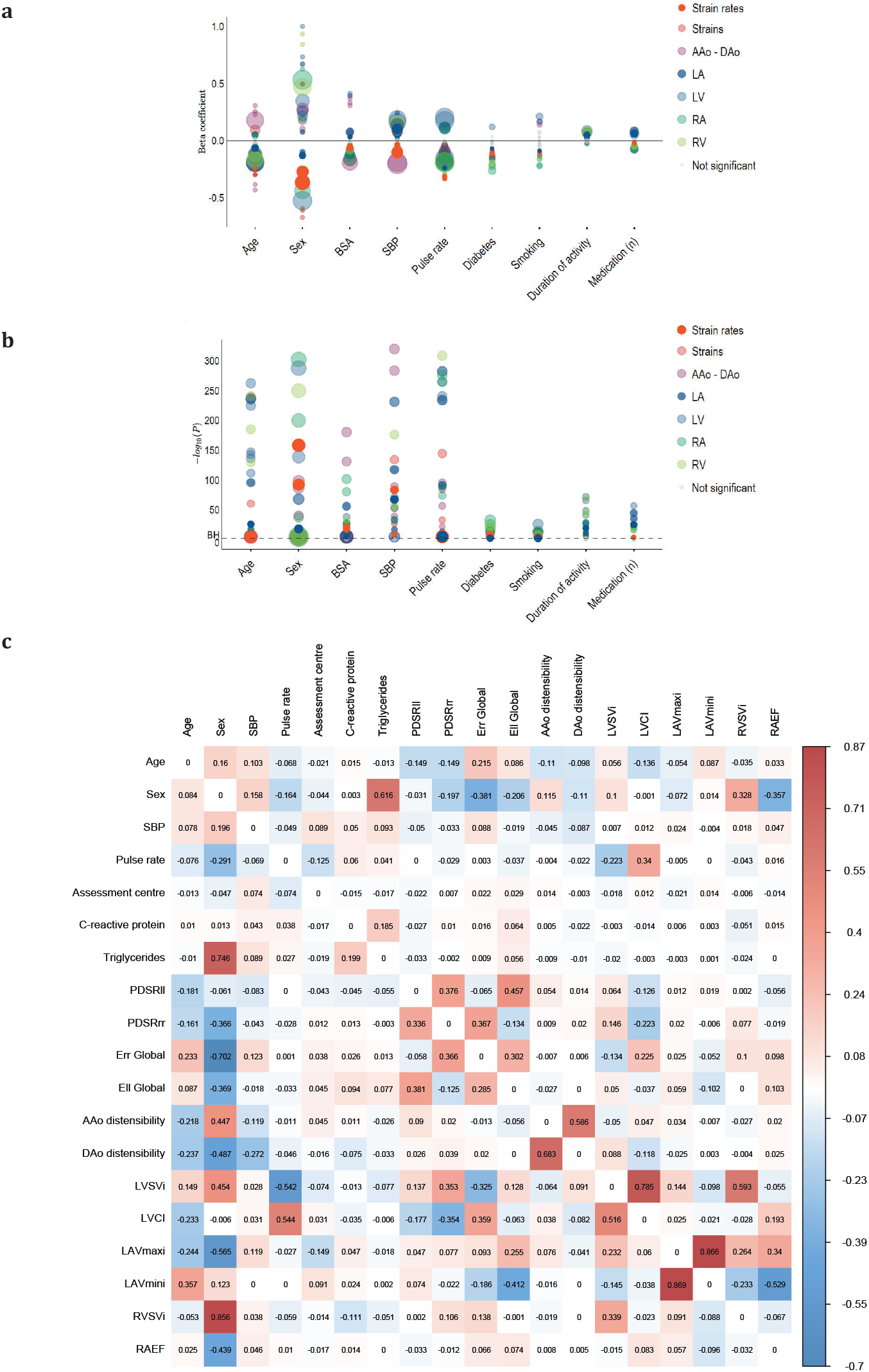
a) Bubble plot showing beta coefficients and b) negative logarithm of the P-values for multiple linear regression analysis between imaging and non-imaging phenotypes. The false discovery rate threshold is shown as a dashed line. c) A plot showing the coefficients for predictors in the LASSO regression model (training set, n=21,403; test set, n=10,217).

**Extended Data Figure 4.**
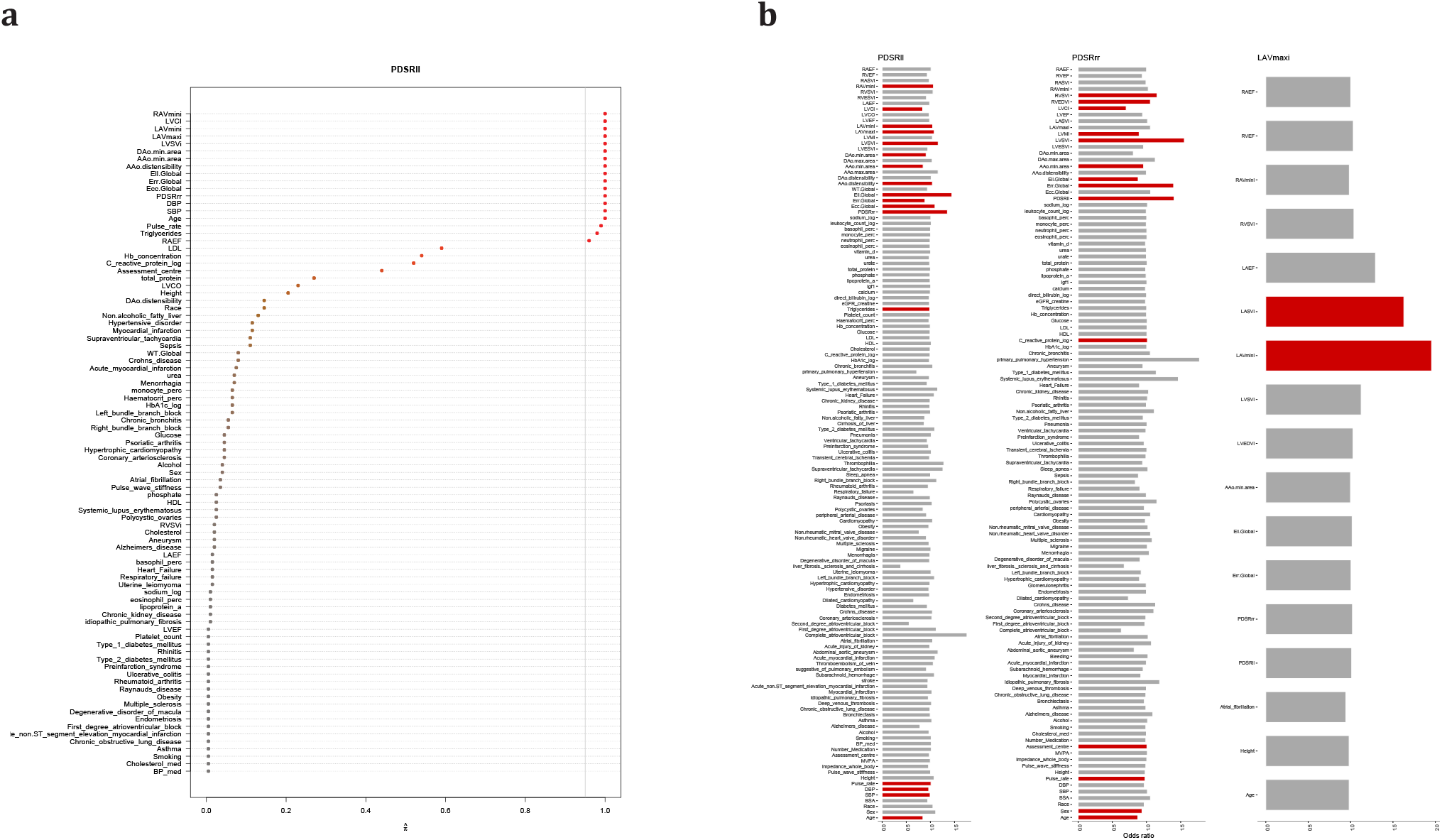
a) Plot showing the covariates selected after stability selection as predictors of PDSR_*ll*_. b) Plot showing the odds ratio of each of the three diastolic function parameters (PDSR_*ll*_, PDSR_*rr*_, and LAVmax_*i*_) with all covariates using LASSO regression and 10-fold cross validation. Red bars indicate variables selected after stability selection.

**Extended Data Figure 5.**
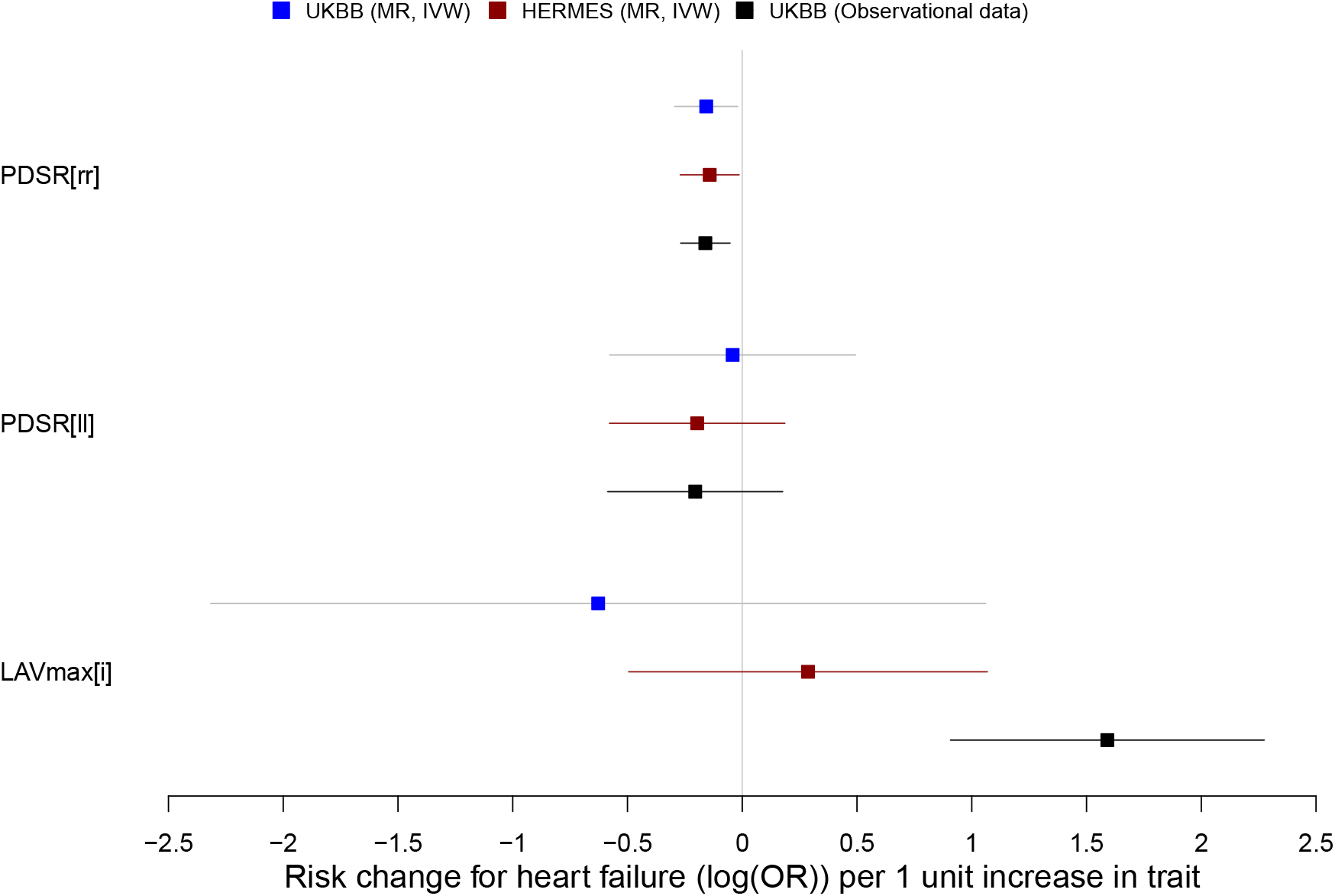
Comparison of association estimates for level of diastolic function traits, PDSR_*rr*_, PDSR_*ll*_ and LAVmax_*i*_, vs. heart failure risk across different approaches (MR approach on UKBB, MR approach using HERMES for the heart failure risk estimates, incident heart failure based on observational data; see Methods for set of considered covariates). Displayed are 95% confidence intervals.

**Extended Data Figure 6.**
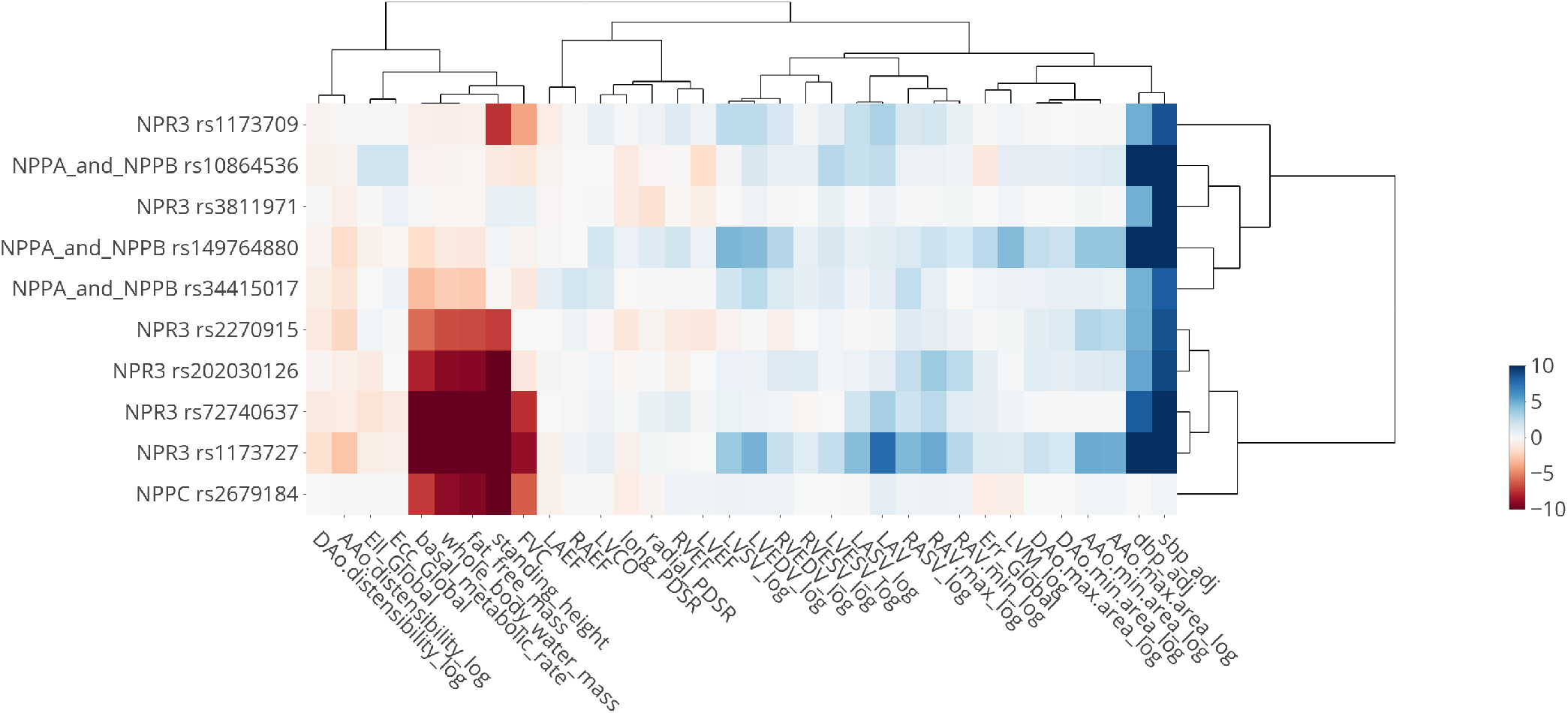
Heatmap of associations with SNPs in genes of the natriuretic peptide pathway. All cardiac imaging traits and traits with a genome-wide significant association with rs1173727 (*NPR3*) were included. SNPs were included if they have a genome-wide significant association with one of these traits except height (height is an extremely polygenic trait with many genome-wide association signals). Values indicate -log10(P-value) of the association test, directionality is aligned to the beta values of the systolic blood pressure (sbp_adj) associations, and to the height associations if there is no significant blood pressure association.

## Extended Data Table

**Extended Data Table 1.** Baseline characteristics of the UK Biobank participants in the study.

| Baseline characteristics | Mean ± SD<br>or n (%) | Cardiac Characteristics from CMR | Mean ± SD<br>or n (%) |
| --- | --- | --- | --- |
| Age (years) | 63.6 ± 7.6 | LV wall thickness (mm) | 5.7 ± 0.8 |
| Sex, men, n (%) | 18,988 (48%) | LV end-diastole volume indexed ($mL/m^2$) | 79 ± 13.8 |
| Race, Nonwhite, n (%) | 1,130 (2.8%) | LV end-systole volume indexed ($mL/m^2$) | 32.1 ± 8.4 |
| Body mass index ($kg/m^2$) | 26.5 ± 4.4 | LV stroke volume indexed ($mL/m^2$) | 46.9 ± 8.4 |
| Body surface area ($m^2$) | 1.9 ± 0.2 | LV ejection fraction (%) | 59.6 ± 6.1 |
| Systolic Blood pressure (mmHg) | 138.2 ± 18.3 | LV cardiac output (ml) | 5.4 ± 1.2 |
| Diastolic blood pressure (mmHg) | 78.6 ± 9.9 | LV cardiac index ($ml/m^2$) | 2.5 ± 0.5 |
| Pulse rate (bpm) | 70 ± 12 | LV mass indexed ($mL/m^2$) | 45.7 ± 8.5 |
| Pulse wave arterial stiffness index (SI) | 9.6 ± 2.9 | LA maximum volume indexed ($ml/m^2$) | 39 ± 11.2 |
| Diabetes mellitus, n (%) | 2,432 (6.2%) | LA minimum volume indexed ($ml/m^2$) | 15.7 ± 7.5 |
| Heart failure, n (%) | 260 (0.66%) | LA stroke volume indexed ($ml/m^2$) | 23.3 ± 5.8 |
| Smoking status |  | LA emptying fraction (%) | 61.2 ± 9.5 |
| Current, n (%) | 1,374 (3.5%) | RV end-diastole volume indexed ($ml/m^2$) | 83.6 ± 15.2 |
| Previous, n (%) | 13,330 (34.1%) | RV end-systole volume indexed ($ml/m^2$) | 35.9 ± 9.3 |
| Never, n (%) | 24,443 (62.4%) | RV stroke volume indexed ($ml/m^2$) | 47.7 ± 8.9 |
| Daily alcohol intake | 6,597 (16.7%) | RV ejection fraction (%) | 57.3 ± 6.1 |
| Duration of physical activity in minutes per day | | RA maximum volume indexed ($ml/m^2$) | 46.4 ± 13.5 |
| Moderate | 53.9 ± 66.2 | RA minimum volume indexed ($ml/m^2$) | 24.7 ± 9.2 |
| Vigorous | 40.3 ± 40.4 | Right atrial stroke volume indexed ($ml/m^2$) | 21.6 ± 6.8 |
| Number of treatment/medications taken | 1.9 ± 2.1 | RA emptying fraction (%) | 47.2 ± 9.5 |
| Blood pressure medication | 2,042 (5.2%) | AAo distensibility indexed ($10^{-3} \cdot mmHg^{-1}$) | 0.97 ± 0.63 |
| Cholesterol medication | 6,015 (15.2%) | AAo maximum area ($mm^2$) | 852.3 ± 188.4 |
| Assessment centre | | AAo minimum area ($mm^2$) | 775.1 ± 183.9 |
| Cheadle | 25,176 (63.6%) | DAo distensibility indexed ($10^{-3} \cdot mmHg^{-1}$) | 1.29 ± 0.8 |
| Reading | 4,361 (11%) | DAo maximum area ($mm^2$) | 476.7 ± 96.8 |
| Newcastle | 10,022 (25.3%) | DAo minimum area ($mm^2$) | 418.1 ± 91.6 |
| Laboratory Biochemical Markers |  | Strains and Strain rates |  |
| HbA1c ($\log(mmole/mol)$) | 3.5 ± 0.13 | Peak diastolic longitudinal strain rates ($PDSR_{ll}, s^{-1}$) | 1.64 ± 0.6 |
| C-reactive protein ($\log(mg/L)$) | 0.13 ± 1.02 | Peak diastolic radial strain rates ($PDSR_{rr}, s^{-1}$) | 5.71 ± 1.9 |
| LDL ($mmol/L$) | 3.6 ± 0.8 | Global circumferential strain ($E_{cc}, \%$) | 22.3 ± 3.4 |
| Glucose ($mmol/L$) | 5.0 ± 0.93 | Global longitudinal strain ($E_{ll}, \%$) | 18.5 ± 2.8 |
| Triglycerides ($\log(mmole/L)$) | 0.36 ± 0.51 | Global radial strain ($E_{rr}, \%$) | 45.1 ± 8.4 |
| eGFR cystatin ($mL \cdot min^{-1} \cdot 1.73 m^{-2}$) | 92 ± 12.2 | | |

**Baseline characteristics and cardiac information from CMR. Values are presented as mean ± SD or n (%).**
\*LV: left ventricle, LA: left atrial, RV: right ventricle, RA: right atrial, AAo: Ascending aortic, DAo: Descending aortic.

