## Supplementary Methods for "Genetic and environmental determinants of diastolic heart function"

**Overview of analysis strategy:** We analysed CMR data from 39,559 participants in UK Biobank using machine learning motion tracking to measure three validated parameters of diastolic function - radial and longitudinal peak end diastolic strain rate ($PDSR_{rr}$ and $PDSR_{ll}$), and maximum indexed atrial volume ($LAVmax_i$). We first explored the associations between diastolic function and environmental factors to define cardiovascular disease risks. We further performed a genome-wide association study of diastolic functional traits and we identified top hits containing genes associated with cardiomyocyte function and cardiomyopathy. Consequences of changes in diastolic function parameters were assessed with polygenic risk scores and mendelian randomization. A summary of our study objectives and conducted analysis is given in Extended Data Fig 1. Further details are provided in the following paragraphs.

**Definition of disease endpoints:** Disease endpoints, i.e. binary traits are defined based on records from general practitioners (GP, available for a subset of the UK Biobank cohort), hospital episode (HES) data and self-reported information. If any of the read2/read3 codes (for GP data), ICD9/ICD10 codes (for HES data), OPC codes (for operational and procedural codes), or any of the self-reported disease codes (UK Biobank field 20002) are reported at least once for each subjects at any time, we consider the subject to be a case and if no code is reported we consider the subject to be a control.

The codes corresponding to the considered disease traits are reported in Supplementary Data 1 (Sheet Disease_definitions). If it says NA, it means that no code of the specific category has been used. Definitions are based on code lists provided by UK Biobank (https://biobank.ctsu.ox.ac.uk/crystal/crystal/auxdata/algorithm_outcome_codes.xlsx), defined by CALIBER clinicians or provided within the CALIBER massive web portal (https://caliberresearch.org/portal/phenotypes).

**Definition and quality control of quantitative phenotypes:** Selected quantitative traits are described in Supplementary Data 1 (Sheet Variable_definitions). Most of the selected traits are directly retrieved from a UK Biobank field which is indicated in the table (column "Field"). Other traits are simple composition of field data and the calculation is described in the column "Details". For most traits, we used the measurement at the first assessment center visit (instance 0). Exceptions are described in the table in the column "Preferred instance". If repeated measurements were considered (e.g. for standing height), the mean value of all observations is used, excluding observations that differ by more than 1.5 standard deviations.

In order to remove extreme outliers, implausible values and to reduce skewness of the data, quantitative traits were systematically cleaned. Decisions on which method was applied to which phenotype were taken after visual inspection. We used the following approaches in the stated ordering (described in column "QC" in the xlsx):

- Exclusion of single, isolated outliers (called outlier in xlsx): we used a step-wise procedure. First, we calculate the median absolute deviation (MAD) and check for each observation (x) the number of other observations (from other subjects) within
$$[x - 0.5 \cdot \mathrm{MAD}, x - 0.5 \cdot \mathrm{MAD}].$$
If less than 30 other observations lie in this interval, we consider the observation to be isolated and set the value x to NA.
- Null deletion (called null in xlsx): remove all values that are 0 (necessary prior to log transformation, sometimes also indicating an implausible value).
- Percentage deletion (called percentage in xlsx): deletes the 0.1% of ordered observations on the top and on the bottom.
- Log-transformation: application of a log-transformation to reduce skeweness.

For the genetic analysis only, we also adjusted blood pressure measurement to the self-reported blood pressure lowering medications. Specifically, we identified subjects with blood pressure lowering medication via UK Biobank field 6177 and 6153 and added 15 mm Hg to the systolic blood pressure and 10 mm Hg to the diastolic blood pressure if blood pressure lowering medications were reported.

**Phenotype Analysis Procedure:** In addition to the above described QC, for the phenotype analysis we used an imputation technique for handling missing values with multiple imputations using predictive mean matching for continuous variables, with 5 imputations and 5 iterations in the R package "mice"[72]. Diagnostic plots were used to test the assumptions underlying linear regression and analysis was performed using the R package "stats".

**LASSO Phenotype Analysis Procedure:** We further fitted a least absolute shrinkage and selection operator (LASSO) model in order to estimate the statistical associations between phenotypes, implemented in the R package "glmnet"[73]. We applied feature selection algorithms using stability selection method that uses resampling to assess the stability of selected phenotypes for a robust selection of covariates using the R package "stabs"[74]. Here, we fitted LASSO models via stability selection procedure setting as predictors the three diastolic function parameters in order to estimate stable statistical associations between the variables. The selection probability "cutoff" was set to 0.95, the per-family error rate was set to 1.0 and the "fitfun" parameter of the "stabsel" function was set as "glmnet.lasso"[75]. Compared to the classical approaches here, the regularized regression approaches provide an alternative way to handle high dimensional data and to identify phenotype associated with the trait of interest using variable selection[76].

We fitted L1-regularized logistic or linear regression (LASSO) that optimises the model coefficient ($\beta$) of the linear regression, where the $\lambda$ parameter represents the strength of the regularization. We adjusted $\lambda$ by a ten-fold cross-validation (CV) method on a training set (68%, n=26,893) including all covariates, using the *cv.glmnet* function from the "glmnet" R package. The lambda.min parameter, which denotes the value that gives minimum mean cross-validated error, was extracted and used for prediction on the test set (32%, n=12,666). The covariates selected from the model selection method were checked for collinearity by computing the variance inflation factor (VIF). Finally, we excluded the covariates with high VIF values (VIF > 5) in order to avoid collinearity. For the visualization of the phenotype interactions we used the R package "corrplot" and for network construction, we used the R package. "circlize"[77].

### UK Biobank genetic data

**Details on quality control** We follow the QC procedure proposed by UK Biobank and exclude subjects with

- Heterozygosity or high missing rate (indicated by field 22027)
- Missmatch between genetic and self-reported sex (indicated by field 22001 and 31)
- Sex chromosome aneuploidy (as indicated by field 22019)
- To exclude subjects which are closely related to others, we use the provided kinship coefficients by UK Biobank generated by the KING software[78]. For each of the pairs of sample with a kinship coefficient >0.884 (i.e. second degree relationship or closer), a single sample was excluded at random.

That leads to 372 subjects from the full genotyped cohort (N=487279) being excluded due to a mismatch between reported and genetically inferred sex, 651 subjects being excluded due to sex chromosomal aneuploidy, 968 subjects being excluded due to a high percentage of missing genotypes and/or heterozygosity rate outliers and 36159 subjects being excluded due to suspected relatedness. This leads to 449263 subjects who passed the genetic QC out of which 36541 subjects were part of the first three data releases of the MRI imaging substudy.

**Ancestry estimation based on reference samples of known ethnicities.** To avoid confounding due to hidden population substructure, we only included individuals forming a well-mixed population in the genetic association tests. As the majority of participants in UK Biobank self-identify as white European, we focused our analysis on this population.

To identify individuals of European ancestry based on their genetics, we compared UK Biobank genotypes to genotypes from the HapMap Phase III study[79] using the R package *plinkQC* (v0.3.3) and following the accompanying vignettes 'Processing HapMap III reference data for ancestry estimation' and 'Ancestry estimation based on reference samples of known ethnicities'[80]. In brief, we downloaded the HapMapIII reference data from ftp://ftp.ncbi.nlm.nih.gov/hapmap/genotypes/2009-01_phaseIII/plink_format/, harmonised and joined the reference with UK Biobank genotypes and conducted principal component analysis. Individuals that clustered with the HapMap III individuals of European ancestry of the PC1/PC2 plot were kept for further analyses. Supplementary Fig 1).

**Sample sizes of study cohorts of genetic analysis:** GWAS analyses were conducted, within the described European cohort that passed genetic QC, in a imaging discovery and imaging replication analysis, in the full imaging cohort and in a non-imaging cohort. The imaging discovery cohort is defined as all samples available within the first two releases of MRI imaging data from UK Biobank (23321 samples after genetic QC) while the replication dataset is defined as all samples from the third release of UK Biobank MRI imaging data (10924 samples after genetic QC). The full imaging dataset is the union of replication and discovery dataset (34242 subjects after genetic QC). The non-imaging cohort is defined as all subjects who have not been included in the first three releases of UK Biobank MRI imaging data (386108 samples after genetic QC). The full European Biobank population consistent of all subjects who passed QC (420365 samples after QC).

**GWAS variant-to-gene mapping and gene annotation:** Supplementary Fig 2 gives an overview of the variant-to-gene mapping. 5 loci reached genome-wide significance ($P = 5 \cdot 10^{-8}$) in the discovery dataset. These were replicated in the replication dataset using a Bonferroni-corrected significance threshold of $\frac{0.05}{5} = 0.01$. An additional 4 loci reached genome-wide significance ($P = 5 \cdot 10^{-8}$) in the full dataset (main text).

The 9 significant loci were mapped to estimated causal genes using the following criteria:

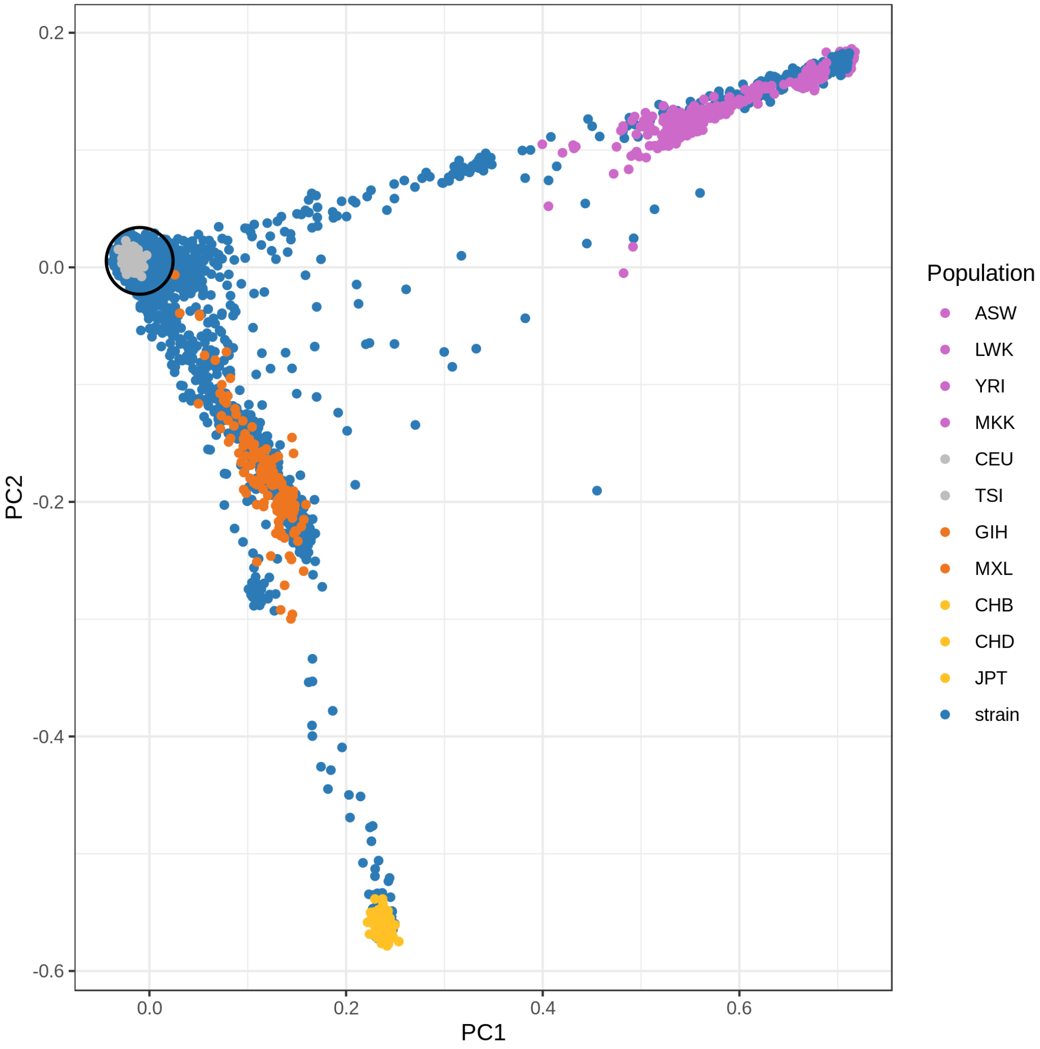


**Supplementary Figure 1. Genetic principal components of UK Biobank and HapMap III reference.** Components 1 and 2 of the principal component analysis on the combined genotypes of the UK Biobank and HapMap III datasets. UK Biobank individuals are depicted in blue, HapMap III individuals colored by their ethnicity. UK Biobank individuals within 1.5 standard deviations distance from the center of the European HapMap individuals (grey) are selected for further analyses.

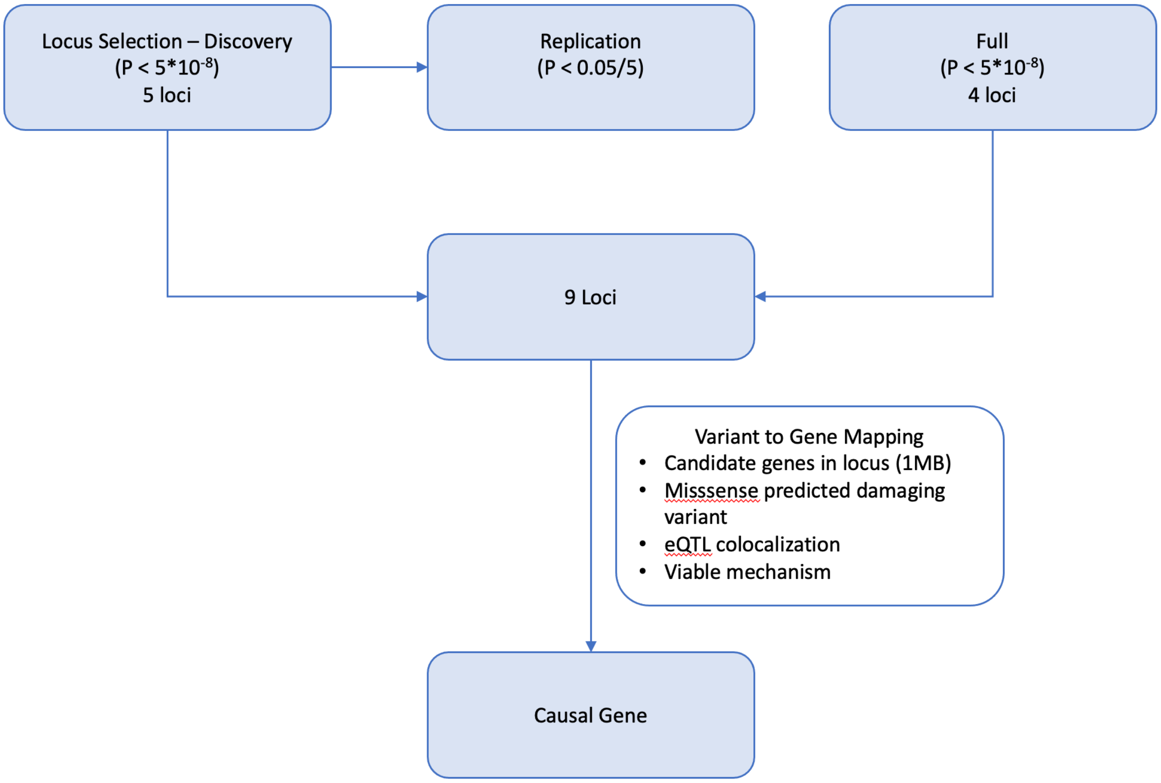


**Supplementary Figure 2. Variant-to-gene mapping.** This flowchart shows the steps taken to go from significant loci (9; 5 from the discovery dataset, and an additional 4 from the full dataset), through to estimated causal genes.

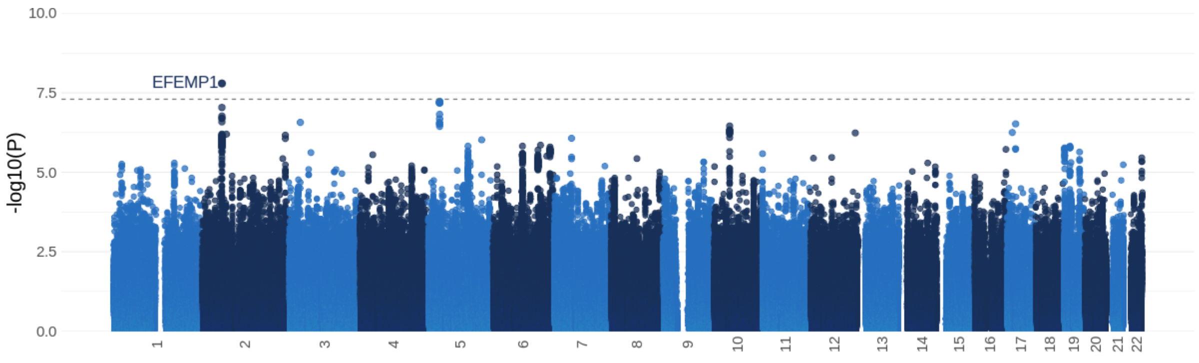


**(a)** LAVmax$_i$

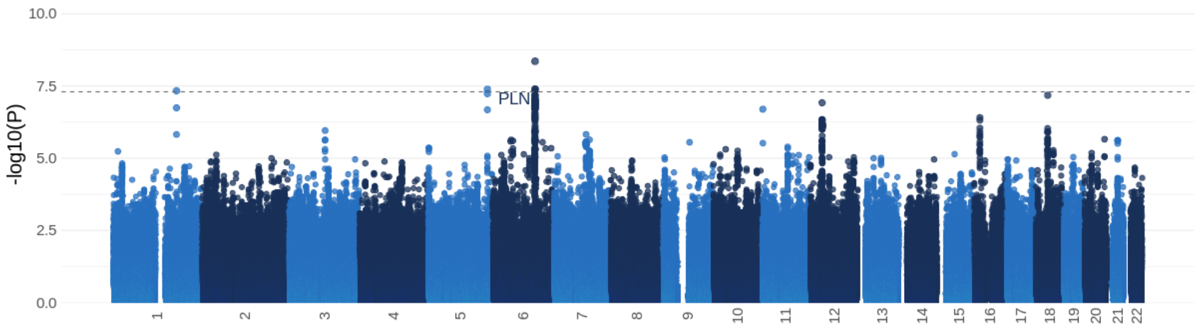


**(b)** PDSR$_{ll}$

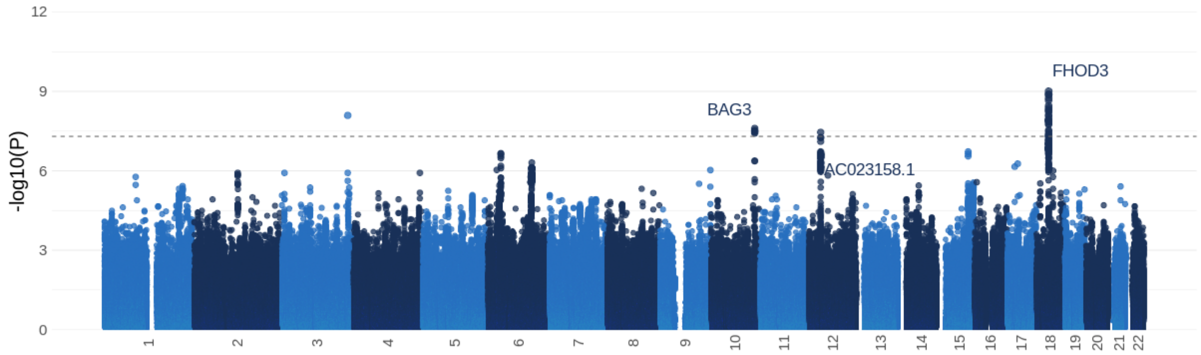


**(c)** PDSR$_{rr}$

**Supplementary Figure 3. Manhattan plots of the GWAS results for diastolic function traits (discovery dataset).** This figure shows the -log10(P-value) on the y axis across all available chromosomal positions (x-axis). The displayed P-value is the BOLT-LMM P-value. The dotted line indicates genome-wide significance ($P = 5 \times 10^{-8}$). Loci that lose genome-wide significance in the full dataset are not annotated.

- Is there evidence that the variant directly affects the function of a candidate gene? Here, VEP annotation[81] was used to test if the variant was predicted damaging or loss of function.

- Is the variant also an eQTL for the candidate gene in a relevant cell type / tissue? The presence of a colocalising e/pQTL was determined by searching the following resources: GTEx, CEDAR, Fairfax 2012, TWINSUK, eQTLGen, GENCORD, HIPSCI, GEUVADIS, Alasso 2018, Nedelec 2016, Blueprint, Quach 2016, Naranbhai 2015, Van de Bunt 2015, and Schwartzentruber 2018. The full summary statistics for the locus were downloaded for possible colocalising eQTLs (i.e. those where the lead variant was also a significant eQTL; $p < 10-3$). Colocalisation was verified or repudiated by a visual inspection of the locus plots.

- Is there any additional functional data linking the variant to the gene? Using https://genetics.opentargets.org/, PCHi-C, DHS-promoter correlation, and enhancer-TSS correlation associations were also leveraged.

- Does the candidate gene's function fit with the GWAS phenotype? Is there supporting evidence from the gene mechanism or additional PheWAS associations, suggesting a viable link between the gene's function and the imaging phenotype?

Using this criteria, the following variant-to-gene mappings were made for the 9 significant loci (see Supplementary Fig 4 for the accompanying colocalisation locus plots):

- **rs2234962** eQTL data and colocalising GWAS strongly suggest that the causal gene here is BAG3. The lead variant, rs2234962, a missense variant found within the BAG3 coding sequence, is also associated with BAG3 expression in blood. The other candidate, MCMBP, has multiple eQTL signals in the same locus but, as can be seen from Supplementary Fig 4, these signals do not colocalise with the radial PDSR signal and are likely driven by a different variant in the locus. **Summary: Effect variant; increases BAG3 expression, increases PDSR$_{rr}$, increases pulse rate, probable causal gene: BAG3, effect independent of pulse rate.**

- **rs2644262** This is an intronic variant for FHOD3 with a colocalising eQTL signal for the gene in heart tissue, but there is also a strong colocalisation with an eQTL for TPGS2 (Supplementary Fig 4). There is a lot of evidence linking FHOD3 to cardiomyopathy (e.g. https://www.targetvalidation.org/target/ENSG00000134775/associations), suggesting FHOD3 is more likely to be the causal gene here. **Summary: Effect variant; increases PDSR$_{rr}$, decreases pulse rate, probable causal gene: FHOD3, effect independent of pulse rate.**

- **rs11970286** This lead variant is closest to PLN, which is strongly associated with cardiomyopathy (https://www.targetvalidation.org/target/ENSG00000198523/associations). An eQTL signal for CEP85L also colocalises with the signal for PDSR$_{ll}$ but literature evidence shows a stronger association between PLN and heart/cardiovascular function. The longitudinal PDSR signal again colocalises with Pulse Rate and Adjusted Diastolic Pulse Rate. **Summary: Effect variant; increases PDSR$_{ll}$, decreases pulse rate, possible causal gene: PLN**

- **rs11170519** For this locus, the probable gene candidates, by location, are AC023158.1, ALG10, AC023158.2, and SYT10. There are eQTLs for AC023158.1 and ALG10 but the colocalisation analysis suggests AC023158.1 (iPSCs and GTEx Nerve Tibial tissue). AC023158.1 is a novel transcript and there are no clear mechanistic links between the other genes and heart phenotypes so the conclusion here is inconclusive. **Summary: Effect variant; increases PDSR$_{ll/rr}$, decreases pulse rate, possible causal gene: inconclusive**

- **rs59985551** rs59985551 is an intronic variant for EFEMP1 and is also associated with increased EFEMP1 expression in Thyroid (GTEx). There are no other eQTLs for genes in this locus (https://genetics.opentargets.org/variant/2_55879793_C_T). EFEMP1 is an extracellular matrix protein mainly associated with eye disease (https://www.targetvalidation.org/target/ENSG00000115380/associations). **Summary: Effect variant; increases LAVmax$_i$, increases blood pressure, possible causal gene: EFEMP1, effect independent of pulse rate.**

- **rs1173727** We find rs1173727 as significant hit from our BOLT-LMM analysis and the most likely variant to gene mapping of rs1173727 shows NPR3 as the best target hit (see https://genetics.opentargets.org/variant/5_32830415_T_C). In line with opentargets.org we assume allele C to be the 'effect' allele. BOLT-LMM thus shows a positive effect on LAVmax$_i$. PheWAS of rs1173727 in UK Biobank shows positive association to diastolic blood pressure, systolic blood pressure, and mean arterial pressure, indicating increased blood pressures for C allele carriers. Colocalisation analysis[82] of NPR3 SNPs from BOLT-LMM compared with diastolic and systolic blood pressure, as well as Mean Arterial Pressure from UK Biobank show high posterior probabilities for colocalisation. PheWAS analysis of binary traits for rs1173727 shows a weak positive association with Heart Failure risk ($P = 6.08 * 10^{-3}$) and a strong association with hypertensive disorder ($P = 1.52 * 10^{-38}$), indicating higher risk for these traits for effect allele carries. Results of

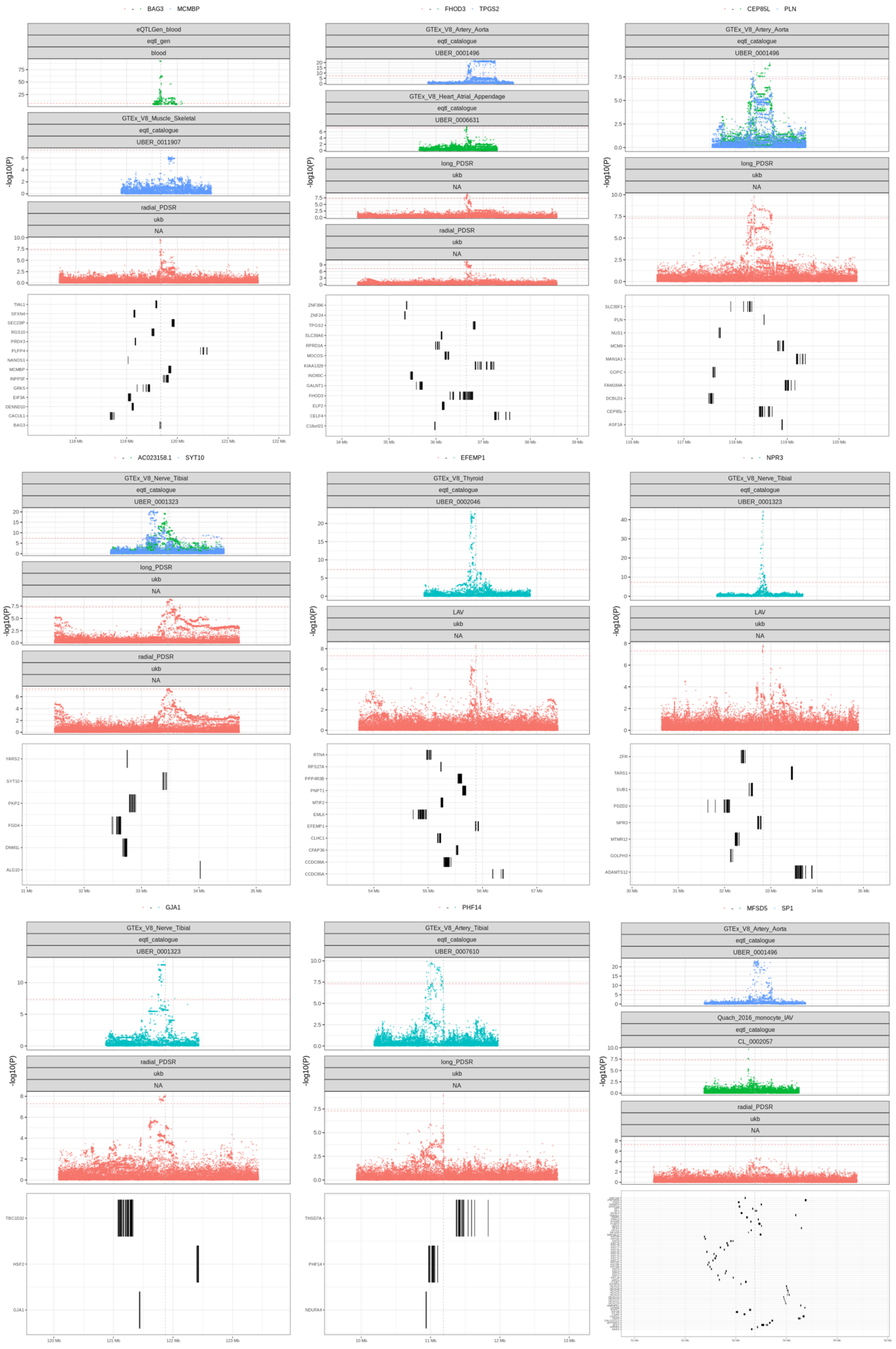


**Supplementary Figure 4. Colocalisation of additional GWAS and eQTL signals at the 9 signifcant loci** For each of the 9 significant loci, the relevant GWAS and eQTL signals of probable causal genes (and some neighboring genes) is shown.

our Mendelian Randomization analysis might indicate a causal connection of $\text{LAVmax}_i$ as a consequence of dbp_adj, i.e. higher LAVmax for higher dpb. Finally, eQTL data for NPR3 show a clean eQTL locus with variants closely located downstream of NPR3. Variant rs1173727 causes an increase in Nerve Tibial, Lung, Adrenal gland and Testis tissues. **Summary: Effect variant; increases NPR3 expression, increases $\text{LAVmax}_i$, increases blood pressure, probable causal gene: NPR3**

- **rs12206253** Although the variant is situated in an intergenic region, there are eQTL associations with nearby genes (HSF2, GJA1, and SERINC1). Supplementary Fig 4 shows that it is only the eQTL for GJA1 that colocalises with the Radial PDSR signal. Additionally, the Radial PDSR signal colocalises with Pulse Rate from UK Biobank. The association with GJA1 is based on "Nerve - Tibial" tissue from GTEx. GJA1 is a gap junction protein that is associated with multiple developmental disorders, including congenital heart disease (https://www.targetvalidation.org/target/ENSG00000152661/associations and https://www.uniprot.org/uniprot/P17302) **Summary: Effect variant; decreases $\text{PDSR}_{rr}$, increases pulse rate, possible causal gene: GJA1**

- **rs10261575** This variant has a strong colocalisation signal with PHF14 down-regulation in blood and PCHi-C evidence linking for the physical interaction between the variant and the gene promoter (https://genetics.opentargets.org/variant/7_11185589_T_C, Supplementary Fig 4). There are no significant PheWAS associations from UK Biobank and PHF14 does not seem to be strongly linked to heart phenotypes by mechanism or additional phenotypes. **Summary: Effect variant; decreases PHF14 expression (blood), increases $\text{PDSR}_{ll}$, possible causal gene: PHF14**

- **rs11170519** This variant has a colocalisation signal with SP1 down-regulation (https://genetics.opentargets.org/variant/12_53374856_C_T, Supplementary Fig 4). However, the locus is gene-dense and it cannot be ascertained if the colocalisation is specific to SP1. SP1 is a zinc finger transcription factor involved in many cellular processes but with no clear link to heart phenotypes. **Summary: Effect variant; decreases SP1 expression, increases $\text{PDSR}_{rr}$, possible causal gene: SP1**

**Supplementary Table 1.** A summary of the lead variant effects on the causal gene expression, the 3 measured diastolic traits, and other heart phenotypes.

| Lead Variant | Causal Gene | Effect on Gene | $\text{LAVmax}_i$ Effect | $\text{PDSR}_{rr}$ Effect | $\text{PDSR}_{ll}$ Effect | Effect on other Heart Phenotypes |
|---|---|---|---|---|---|---|
| rs2234962 | *BAG3* | **↑** | ↑ | **↑** | ↑ | **↓** pulse rate |
| rs2644262 | *FHOD3* | Unknown | ↑ | **↑** | **↑** | **↓** pulse rate |
| rs11970286 | *PLN* | Unknown | ↑ | ↑ | **↑** | **↓** pulse rate |
| rs1580396 | *AC023158.1* | **↓** | ↑ | **↑** | **↑** | **↓** pulse rate |
| rs59985551 | *EFEMP1* | **↑** | **↑** | ↑ | ↑ | **↑** blood pressure |
| rs1173727 | *NPR3* | **↑** | **↑** | ↓ | ↑ | **↑** blood pressure |
| rs12206253 | *GJA1* | **↑** | ↓ | **↓** | ↓ | **↑** pulse rate |
| rs10261575 | *PHF14* | **↓** | ↑ | ↑ | **↑** | None |
| rs11170519 | *SP1* | **↓** | ↑ | **↑** | ↑ | None |

**Details on Mendelian randomization analysis:** Mendelian randomization (MR) can be used to infer causal relationships between risk factors and outcomes. However, unbiased results are only expected if the assumption of MR methods hold Supplementary Fig 5. These assumptions focus on the following three aspects

- The instrumental variables, i.e., the SNPs are associated with the risk factor (also called exposure).
- The SNPs are independent of any potential confounder of the relationship between the considered risk factor and outcomes.
- The SNPs influence the outcome only via the risk factor (the exposure), i.e. there is no other causal path between the SNP and the outcome apart from the path via the risk factor.

Certainly, these assumptions cannot be verified numerically. That is why we use multiple MR methods and conduct additional sensitivity analysis as described in the following sections.

**Selection of considered MR methods** Our main analysis is based on estimates obatined from the inverse-variance weigted (IVW) method. If the assumptions of MR analysis are valid, this methods is well-powered and provides unbiased causal

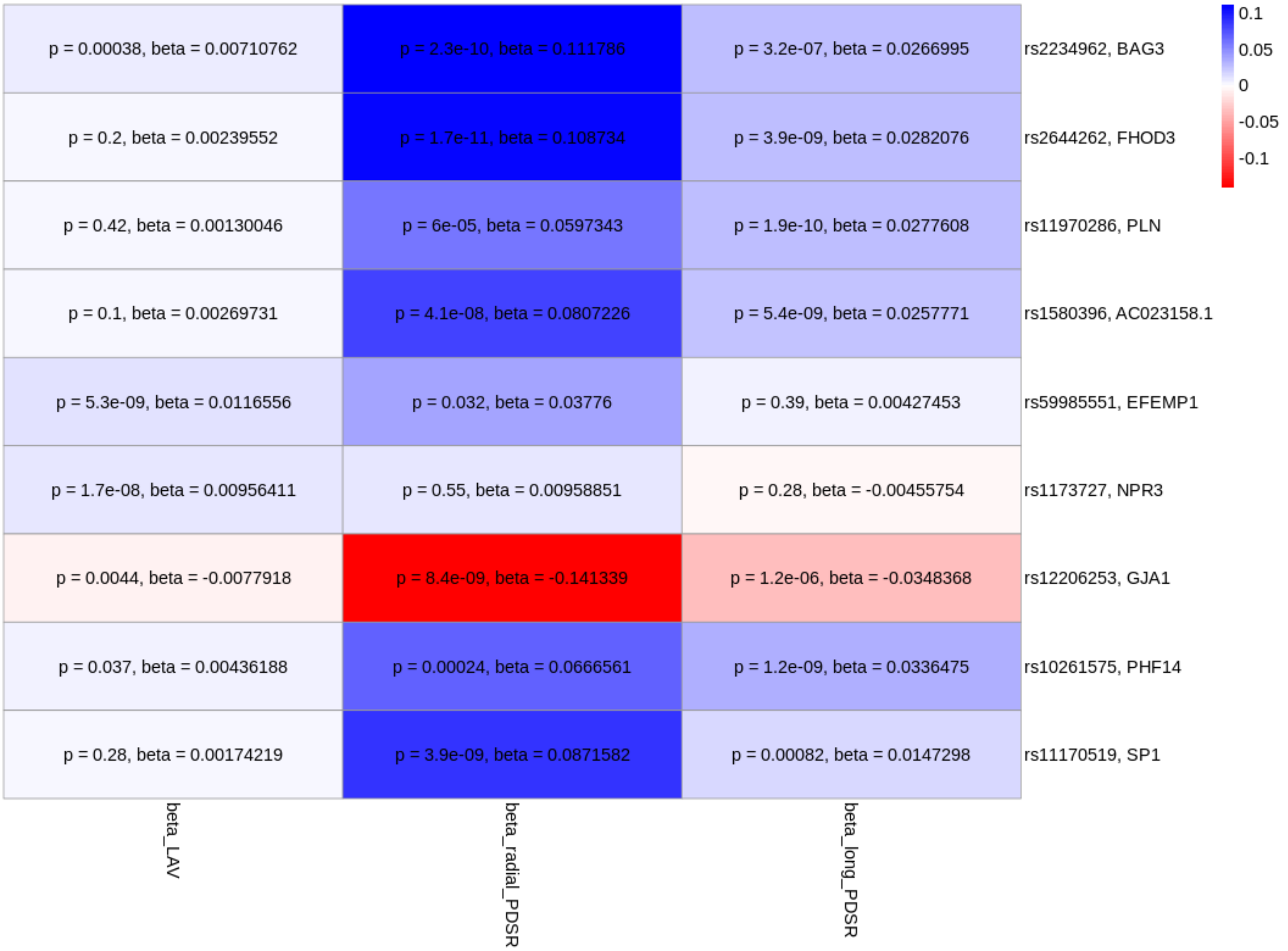


**Supplementary Figure 5. Heatmap of Variant Effects** The lead variant effect size and p-values for each of the diastolic traits.

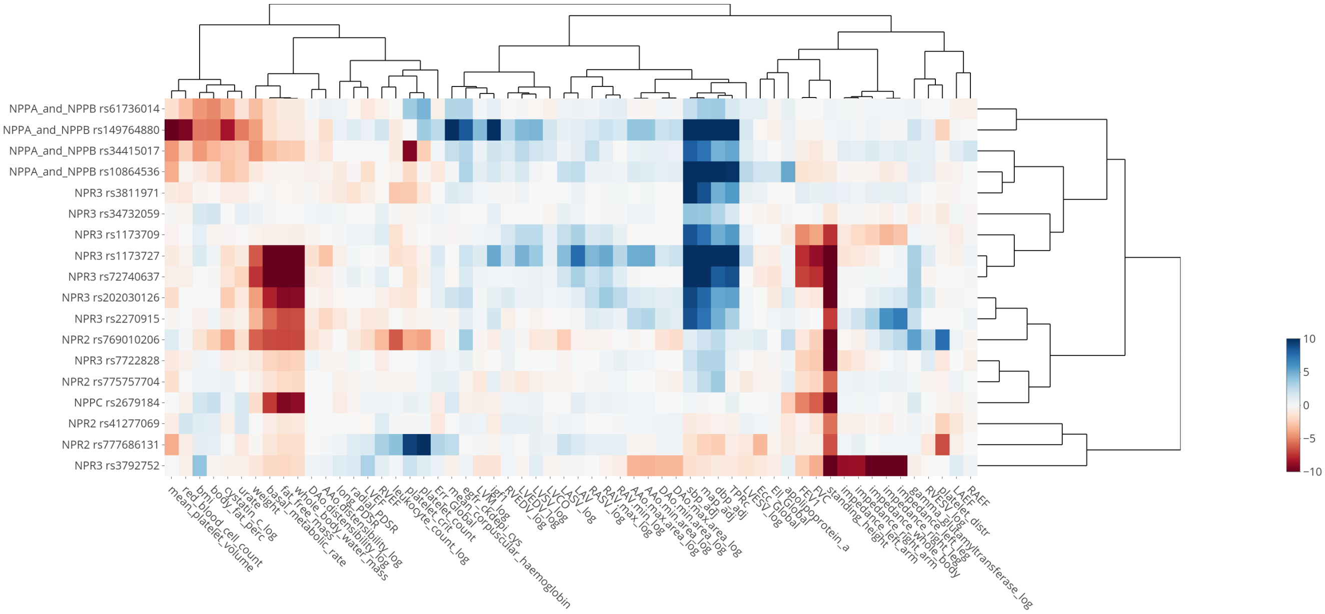


**Supplementary Figure 6. Heatmap of associations with SNPs in genes of the natriuretic peptide pathway** All cardiac imaging traits and available quantitative traits from UK Biobank with a genome-wide significant association with rs1173727 were included. SNPs were included if they have a suggestive association with one of these traits. Values indicate -log10(P-value) of the association test, directionality is aligned to the $\beta$ values of the systolic blood pressure (sbp_adj) associations, and to the height associations if there is no significant blood pressure association.

estimates. However, since it is not possible to confirm the assumptions in a data-driven way, we also consider additional MR approaches to increase our confidence in the identified causal relationships: the weighted median method, MR-Egger and Mendelian Randomization Pleiotropy RESidual Sum and Outlier (MR-PRESSO). The median based method has a higher robustness against invalid instrument bias (here: selection of SNPs which do not fulfill the MR assumptions) and still provides consistent estimates if at least half of the selected SNPs are valid instruments. MR-Egger is a method that can be used for detecting pleiotropy as well as obtaining precise causal estimates in scenarios with pleiotropic instruments. In contrast, the power of MR-Egger compared to IVW is lower if the MR assumptions are valid, leading to less precise estimates. Last, we consider MR-PRESSO which tests explicitly for outliers which could indicate pleiotropic SNPs.

**Details of workflow** Since we investigate a fairly high number of potential causal relationship within this analysis, we needed to establish a workflow that we can use for identifying interesting associations for manual sensitivity analysis. The input for our analysis are the GWAS results obtained from PLINK (for binary traits) or BOLT-LMM (for quantitative traits). Within our analysis, we use the following steps as shown in the flow chart in Supplementary Fig 7:

- Identify overlap in SNP sets between trait 1 and trait 2 GWAS results (snpset).
- Perform clumping on trait 1 GWAS, restricted to the identified snpset from the previous step. Parameters for clumping are set to $R^2 = 0.1$ (in a window of 1000kb) and considering all SNPs with $P < 10^{-6}$. The LD structure from the full QCed European UK Biobank population is used. Clumping was performed with PLINK1.9.
- We extract point estimates and standard deviations for the selected SNPs both from trait 1 GWAS and trait 2 GWAS and use these as the input for the MR analyses. IVW, MR-Egger and the weighted median approach are performed with `MendelianRandomization` R-package[83]. MR-PRESSO is performed with the `MRPRESSO` R-package[84]. For the MR-PRESSO analysis, the hyperparameter NbDistribution is set to 5000 and the SignifThreshold parameter is set to 0.05.
- We judge an association to be "potentially causal" if the IVW analysis leads to a $P < 0.01$, at least two of the sensitivity analysis confirm the results with at least suggestive significance ($P < 0.05$) and none of the other three methodologies

show conflicting results in terms of directionality. For example, if the IVW indicates a positive causal association, none of the other three methods should have 90 % confidence interval that fully falls in the negative range.

- If a potential causal association is found, we perform the supplementary sensitivity analysis as described in the section below if feasible.
- All steps are repeated starting with the clumping on the trait 2 GWAS for the hypothesis of a reverse causal association.

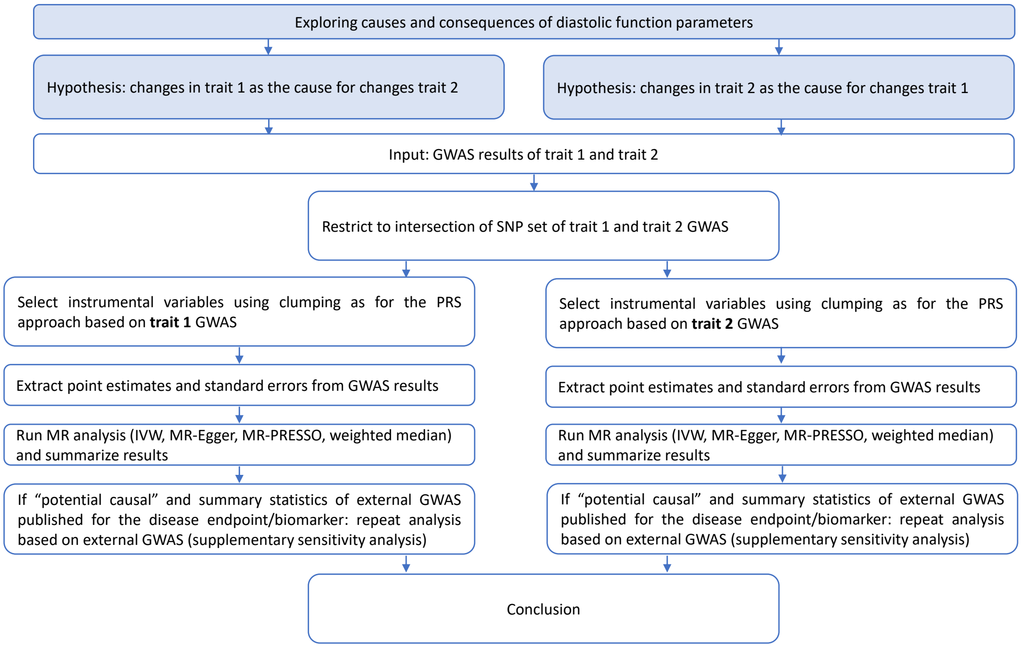


**Supplementary Figure 7.** Flowchart explaining the analysis strategy for the MR approaches. Rules for decision making and details of hyperparameters (e.g., for clumping) are described in the Method section and/or in the Supplementary material.

**Supplementary sensitivity analysis for MR analysis:** Supplementary sensitivity analysis were only conducted for associations which were judge as "potential causal" with the described workflow. First, we explored the exclusion of SNPs with potential pleitropic effects as identified with the HOPS method[85] by making use of their published GWAS results. However, we note a substantial loss in power due to the limited overlap of the SNP sets and therefore do not show any results here.

As a second supplementary sensitivity analysis, we replaced for the non-diastolic traits our own UK Biobank GWAS with published GWAS from large-scale consortia (if available). This can serve as an independent validation and check of robustness of the results. If the sample size of the published GWAS is larger than UK Biobank, this may increase the power of the study with the disadvantage that we do not have full control on the definition of the phenotype, potential heterogeneity between included studies and the details of the conducted analysis. In addition, since UK Biobank is typically part of these GWAS (if the sample size is larger than UK Biobank), an analysis based on published summary statistics is not a full two-sample approach since subjects which are part of the imaging traits GWAS are also part of the GWAS for the other trait of interest.

The MR analysis with the external summary statistics are conducted in a similar way to the main analysis described in this paper: we first intersect the SNP sets between the GWAS for the pair of traits (trait 1 and trait 2). If the effect alleles to not match, we switch the signs of the effect estimates. Afterwards, we run IVW MR analysis, MR-Egger, median-based MR analysis and MR-PRESSO. Note that the restriction of the candidate SNP sets to the overlapping sets between our GWAS and the external GWAS may reduce the power of this analysis in the case that the SNP sets are not widely overlapping and, for example, the top hits for the trait 1 GWAS are missing in the external GWAS.

In a systematic search, we identified the HERMES consortium meta-analysis of heart failure GWAS as a reasonable candidate for this sensitivity analysis. They have investigated heart failure association results across 26 GWAS studies. These studies consists in total of 47.309 cases and 930.014 controls and are case-control studies or population-based studies. Note that UK Biobank is a population-based study and also included within HERMES. Full details on the included studies and the set-up of the meta-analysis can be found in the respective publication[86].

### Supplementary Results

**Univariable and Multivariable Associations:** A summary of the phenotype association analysis is given in Extended Data Fig 3. The associations between phenotypes were analysed using both univariable (Supplementary Table 2 and 3) and multivariable linear regression models (Extended Data Fig 4), to explore the relationship between each diastolic function parameter and cardiovascular risk factors.

**Supplementary Table 2.** Univariable associations between each imaging phenotypes and each non-imaging phenotypes.

| | $PDSR_{ll}$ | $PDSR_{rr}$ | $E_{cc}$ Global | $E_{rr}$ Global | $E_{ll}$ Global |
|---|---|---|---|---|---|
| Age | -0.33 (-0.34,-0.32)* | -0.26 (-0.27,-0.25)* | 0.01 ($-2\cdot10^{-3}$,0.02) | 0.11 (0.10,0.12)* | -0.02 (-0.03,-0.01)* |
| Sex | -0.34 (-0.36,-0.33)* | -0.40 (-0.42,-0.38)* | -0.67 (-0.68,-0.65)* | -0.64 (-0.66,-0.62)* | -0.48 (-0.50,-0.46)* |
| BSA | -0.13 (-0.14,-0.13)* | -0.17 (-0.18,-0.16)* | -0.26 (-0.27,-0.25)* | -0.22 (-0.23,-0.21)* | -0.13 (-0.14,-0.12)* |
| SBP | -0.22 (-0.23,-0.22)* | -0.15 (-0.16,-0.14)* | $8\cdot10^{-4}(-9\cdot10^{-3},0.01)$ | 0.11 (0.10,0.12)* | -0.03 (-0.04,-0.02)* |
| DBP | -0.22 (-0.23,-0.21)* | -0.20 (-0.21,-0.19)* | -0.11 (-0.12,-0.10)* | -0.04 (-0.05,-0.03)* | -0.10 (-0.11,-0,09)* |
| Pulse rate | -0.26 (-0.27,-0.25)* | -0.28 (-0.29,-0.27)* | -0.02 (-0.03,-0.01)* | 0.04 (0.03,0.05)* | -0.10 (-0.11,-0.09)* |
| Diabetes | -0.38 (-0.42,-0.33)* | -0.37 (-0.42,-0.32)* | -0.21 (-0.25,-0.17)* | -0.07 (-0.11,-0.03)* | -0.19 (-0.23,-0.15)* |
| Smoking | -0.02 (-0.06,0.02) | -0.05 (-0.09,-0.01)* | -0.10 (-0.14,-0.06)* | -0.13 (-0.17,-0.09)* | -0.13 (-0.17,-0.09)* |
| Activity | 0.02 (0.01,0.03)* | 0.03 (0.02,0.04)* | -0.02 (-0.03,-0.01)* | -0.02 (-0.03,-0.01) | $-7\cdot10^{-3}(-0.02,3\cdot10^{-3})$ |
| Medication | -0.12 (-0.13,-0.11)* | -0.12 (-0.13,-0.12)* | -0.04 (-0.05,-0.03)* | 0.01 ($-3\cdot10^{-3}$,0.05) | -0.04 (-0.05,-0.02)* |
| | AAo distensibility | DAo distensibility | AAo max area | AAo min area | DAo max area |
| Age | -0.47 (-0.47,-0.46)* | -0.43 (-0.44,-0.42)* | 0.19 (0.18,0.20)* | 0.27 (0.26,0.28)* | 0.25 (0.24,0.26)* |
| Sex | -0.14 (-0.16,-0.12)* | -0.24 (-0.26,-0.22)* | 0.79 (0.77,0.81)* | 0.75 (0.73,0.77)* | 1.10 (1.09,1.12)* |
| BSA | -0.10 (-0.11,-0.09)* | -0.16 (-0.17,-0.15)* | 0.42 (0.41,0.43)* | 0.39 (0.38,0.40)* | 0.55 (0.55,0.56)* |
| SBP | -0.33 (-0.34,-0.32)* | -0.35 (-0.36,-0.34)* | 0.20 (0.20,0.21)* | 0.23 (0.22,0.24)* | 0.25 (0.24,0.26)* |
| DBP | -0.16 (-0.17,-0.15)* | -0.17 (-0.18,-0.16)* | 0.24 (0.23,0.25)* | 0.24 (0.23,0.25)* | 0.23 (0.22,0.24)* |
| Pulse rate | -0.06 (-0.07,-0.05)* | -0.08(-0.09,-0.07)* | -0.10 (-0.11,-0.09)* | -0.07 (-0.08,-0.06)* | -0.16 (-0.16,-0.15)* |
| Diabetes | -0.2 (-0.24, -0.16)* | -0.26 (-0.30,-0.22)* | 0.11 (0.07,0.15)* | 0.12 (0.08,0.16)* | 0.17 (0.13,0.21)* |
| Smoking | 0.12 (0.07,0.16)* | 0.13 (0.09,0.17)* | 0.03 (0.02,0.07) | $2\cdot10^{-3}(-0.04,0.04)$ | 0.08 (0.04,0.12)* |
| Activity | 0.01 ($3\cdot10^{-4}$,0.02)* | 0.02 (0.01,0.03)* | 0.03 (0.01,0.03)* | 0.03 (0.02,0.04)* | 0.04 (0.03,0.05)* |
| Medication | -0.12 (-0.13,-0.11)* | -0.14 (-0.15,-0.13)* | 0.09 (0.08,0.10)* | 0.10 (0.09,0.11)* | 0.10 (0.09,0.11)* |
| | DAo min area | LVWT | LVEDVi | LVESVi | LVSVi |
| Age | 0.33 (0.32,0.34)* | 0.09 (0.08,0.10)* | -0.13 (-0.14,-0.12)* | -0.08 (-0.9,-0.07)* | -0.13 (-0.14,-0.12)* |
| Sex | 1.08 (1.06,1.10)* | 1.28 (1.26,1.30)* | 0.70 (0.67,0.71)* | 0.77 (0.75,0.79)* | 0.36 (0.34,0.38)* |
| BSA | 0.52 (0.51,0.53)* | 0.66 (0.65,0.67)* | 0.20 (0.18,0.20)* | 0.24 (0.23,0.25)* | 0.07 (0.06,0.08)* |
| SBP | 0.29 (0.28,0.30)* | 0.36 (0.35,0.37)* | 0.05 (0.04,0.06)* | 0.03 (0.02,0.04)* | 0.05 (0.04,0.06)* |
| DBP | 0.24 (0.23,0.25)* | 0.32 (0.31,0.33)* | -0.02 (-0.03,-0.01)* | 0.03 (0.02,0.04)* | -0.07 (-0.08,-0.06)* |
| Pulse rate | -0.11 (-0.12,-0.10)* | -0.06 (-0.07,-0.05)* | -0.34 (-0.34,-0.33)* | -0.22 (-0.23,-0.21)* | -0.33 (-0.34,-0.32)* |
| Diabetes | 0.21 (0.17, 0.25)* | 0.54 (0.50,0.58)* | -0.21 (-0.25,-0.17)* | -0.06 (-0.10,-0.02)* | -0.29 (-0.33,-0.25)* |
| Smoking | 0.05 (0.01,0.09)* | 0.17 (0.13,0.20)* | 0.06 (0.02,0.10)* | 0.08 (0.04,0.12)* | -0.02 (-0.02,0.05) |
| Activity | 0.03 (0.02,0.04)* | 0.02 (0.01,0.03) | 0.13 (0.12,0.14)* | 0.1 (0.09,0.11)* | 0.11 (0.10,0.12)* |
| Medication | 0.12 (0.11,0.13)* | 0.15 (0.14,0.16)* | -0.07 (-0.08,-0.06)* | -0.03 (-0.04,-0.02)* | -0.09 (-0.10,-0.08)* |
| | LVEF | LVCO | LVCI | LVMi | LAVmaxi |
| Age | 0.01 ($2\cdot10^{-3}$,0.02)* | -0.11 (-0.12,-0.10)* | -0.08 (-0.09,-0.07)* | 0.01 ($-3\cdot10^{-3}$,0.02) | -0.02 (-0.03,-0.01)* |
| Sex | -0.56 (-0.57,-0.54)* | 0.73 (0.71,0.74)* | 0.13 (0.11,0.15)* | 1.23 (1.21,1.25)* | 0.03 (0.01,0.05) |
| BSA | -0.21 (-0.22,-0.20)* | 0.51 (0.50,0.52)* | 0.03 (0.02,0.04)* | 0.45 (0.45,0.46)* | 0.03 (0.02,0.04)* |
| SBP | 0.02 (0.01,0.03)* | 0.20 (0.19,0.21)* | 0.15 (0.14,0.16)* | 0.30 (0.29,0.31)* | 0.08 (0.07,0.09)* |
| DBP | -0.08 (-0.09,-0.07)* | 0.21 (0.20,0.22)* | 0.10 (0.09,0.11)* | 0.21 (0.20,0.22)* | -0.03 (-0.04,-0.02)* |
| Pulse rate | $2\cdot10^{-3}(-8\cdot10^{-3},0.01)$ | 0.15 (0.14,0.16)* | 0.19 (0.18,0.20)* | -0.22 (-0.24,-0.22)* | -0.22 (-0.23,-0.21)* |
| Diabetes | -0.13 (-0.17,-0.09)* | 0.25 (0.21, 0.29)* | -0.02 (-0.02,0.06) | 0.21 (0.17,0.3)* | -0.07 (-0.12,-0.02)* |
| Smoking | -0.08 (-0.12,-0.04)* | 0.09 (0.05,0.13)* | 0.05 (0.01,0.09) | 0.15 (0.11,0.19)* | -0.05 (-0.10,-0.01)* |
| Activity | -0.03 (-0.04,-0.02)* | $2\cdot10^{-3}(-0.01,0.01)$ | 0.03 (0.02,0.04)* | 0.10 (0.09,0.11)* | 0.06 (0.05,0.07)* |
| Medication | -0.03 (-0.04,-0.02)* | 0.04 (0.03,0.05)* | -0.02 (-0.03,-0.01)* | 0.05 (0.04,0.06)* | 0.04 (0.03,0.05)* |
| | LAVmini | LASVi | LAEF | RVEDVi | RVESVi |
| Age | 0.09 (0.08,0.09)* | -0.15 (-0.16,-0.14)* | -0.13 (-0.14,-0.12)* | -0.14 (-0.15,-0.13)* | -0.12 (-0.13,-0.11)* |
| Sex | 0.11 (0.09,0.13)* | -0.09 (-0.11,-0.07)* | -0.15 (-0.17,-0.13)* | 0.86 (0.84,0.88)* | 0.97 (0.95,0.99)* |
| BSA | 0.08 (0.07,0.09)* | -0.04 (-0.05,-0.03)* | -0.10 (-0.11,-0.09)* | 0.25 (0.24,0.26)* | 0.32 (0.31,0.33)* |
| SBP | 0.07 (0.06,0.08)* | 0.05 (0.04,0.06)* | -0.03 (-0.04,-0.02)* | $2\cdot10^{-3}(-0.01,0.01)$ | -0.05 (-0.06,-0.04)* |
| DBP | -0.01 (-0.02,$-2\cdot10^{-3}$)* | -0.04 (-0.05,-0.03)* | 0.02 (0.01,0.03)* | -0.01 (-0.02,$1\cdot10^{-3}$) | 0.02 (0.01,0.03)* |
| Pulse rate | -0.18 (-0.19,-0.17)* | -0.19 (-0.20,-0.18)* | 0.11 (0.10,0.12)* | -0.34 (-0.35,-0.33)* | -0.26 (-0.27,-0.25)* |
| Diabetes | 0.07 (0.03,0.10)* | -0.21 (-0.25, -0.17)* | -0.16 (-0.20,-0.12)* | -0.27 (-0.31,-0.23)* | -0.13 (-0.17,-0.09)* |
| Smoking | -0.04 (-0.18,$-4\cdot10^{-3}$)* | -0.04 (-0.08,$-5\cdot10^{-3}$)* | $-4\cdot10^{-4}(-0.04,0.04)$ | 0.02 (0.02,0.06) | 0.06 (0.02,0.10) |
| Activity | 0.04 (0.03,0.05)* | 0.08 (0.07,0.09)* | -0.01 (-0.02,$-2\cdot10^{-3}$) | 0.13 (0.12,0.14)* | 0.10 (0.10,0.11)* |
| Medication | 0.10 (0.09,0.11)* | -0.06 (-0.07,-0.05)* | -0.11 (-0.13,-0.10)* | -0.11 (-0.12,-0.10)* | -0.08 (-0.09,-0.07)* |

**LASSO Phenotype Analysis:** Here we fitted the least absolute shrinkage and selection operator in order to estimate the statistical associations between phenotypes and evaluated all three model selection methods, however we chose the stability selection method for its robust selection providing a total of 26 optimal variables. A plot showing the stability selection setting as predictor the $PDSR_{ll}$ and the odds ration quantifying the strength of the associations between each of the three diastolic function parameters and all covariates, employed with the LASSO regression method, is given in (Extended Data Fig 4), where the red bars indicate the variables selected after the stability selection method. We then ran regression diagnostics on the model with the selected variables, to exclude a possible collinearity inappropriately influencing our multivariate models and we included 12 imaging consisting of one imaging phenotype of each of the four cardiac chambers (LV, LA, RV, RA), one of the

**Supplementary Table 3.** Univariable associations between each imaging phenotypes and each non-imaging phenotypes (continued from Supplementary Table 2).

| | RVSVi | RVEF | RAVmaxi | RAVmini | RASVi |
|---|---|---|---|---|---|
| Age | -0.12 (-0.13,-0.11)* | 0.03 (0.02,0.04)* | 0.04 (0.03,0.05)* | 0.05 (0.04,0.06)* | 0.01 ($3\cdot10^{-4}$,0.02) |
| Sex | 0.46 (0.44,0.48)* | -0.66 (-0.68,-0.64)* | 0.40 (0.38,0.42)* | 0.56 (0.54,0.58)* | 0.02 ($3\cdot10^{-4}$,0.04) |
| BSA | 0.10 (0.09,0.11)* | -0.26 (-0.27,-0.25)* | 0.01 ($-5\cdot10^{-4}$,0.02) | 0.10 (0.09,0.11)* | -0.11 (-0.12,-0.10)* |
| SBP | 0.06 (0.05,0.07)* | 0.09 (0.08,0.10)* | -0.04 (-0.05,-0.03)* | -0.04 (-0.06,-0.04)* | -0.02 (-0.03,-0.01)* |
| DBP | -0.04 (-0.05,-0.03)* | -0.05 (-0.06,-0.04)* | -0.07 (-0.08,-0.06)* | -0.05 (-0.05,-0.03)* | -0.06 (-0.07,-0,05)* |
| Pulse rate | -0.31 (-0.32,-0.31)* | 0.03 (0.02,0.04)* | -0.24 (-0.25,-0.23)* | -0.25 (-0.26,-0.24)* | -0.13 (-0.14,-0.12)* |
| Diabetes | -0.33 (-0.37,-0.29)* | -0.13 (-0.17, -0.09)* | -0.36 (-0.40,-0.32)* | -0.27 (-0.31,-0.23)* | -0.34 (-0.38,-0.30)* |
| Smoking | -0.03 (-0.06,-0.02) | -0.07 (-0.11,-0.03)* | -0.09 (-0.13,-0.05)* | -0.05 (-0.09,-0.01)* | -0.10 (-0.14,-0.06)* |
| Activity | 0.11 (0.10,0.12)* | -0.02 (-0.03,-0.01)* | 0.10 (0.09,0.11)* | 0.10 (0.09,0.11)* | 0.07 (0.05,0.08)* |
| Medication | -0.11 (-0.12,-0.10)* | -0.01 (-0.02,$1\cdot10^{-4}$) | -0.09 (-0.10,-0.08)* | -0.06 (-0.07,-0.05)* | -0.09 (-0.10,-0.08)* |

| | RAEF |
|---|---|
| Age | -0.02 (-0.03,-0.01)* |
| Sex | -0.52 (-0.54,-0.50)* |
| BSA | -0.18 (-0.19,-0.17)* |
| SBP | 0.04 (0.03,0.05)* |
| DBP | $5\cdot10^{-3}(-5\cdot10^{-3},0.02)$ |
| Pulse rate | 0.14 (0.13,0.15)* |
| Diabetes | $4\cdot10^{-4}(-0.04,0.04)$ |
| Smoking | -0.04 (-0.08,$1.3\cdot10^{-3}$) |
| Activity | -0.05 (-0.06,-0.04)* |
| Medication | $1.1\cdot10^{-3}(-0.01,0.01)$ |

n = 39,559 subjects were analysed with available information for all independent variables. The values are depicted as beta coefficient (95% $CI$). Independent variables include age, sex, body surface area (BSA), systolic blood pressure (SBP), diastolic blood pressure (DBP), pulse rate, diabetes, smoking status, duration of moderate to vigorous physical activity (MVPA) and number of medication or treatment taken. For continuous variables, the coefficient describes the effect of the variable. For binary variables, the coefficient describes the effect with a change in the variable from 0 to 1. * $P < 0.05$.

relevant strains (Ecc, Err, Ell) and two aortic sections (Aao, Dao) where possible and 7 non-imaging phenotypes in order to avoid collinearity, in our final multivariable model.

**Comparison of distribution of PRS within MRI and non-MRI subset** Supplementary Fig 8 shows the distribution of individual subject PRS for the MRI subset (subjects who were part of the first three releases of UKBB imaging data) vs. the non-MRI subset (subjects who were not part of the first three releases of UKBB imaging data). This indicates that there is no systematic bias in the genetic architecture of the traits of interest.

**Results of supplementary sensitivity analysis for MR approaches:** In this part, we briefly describe the results from the supplementary sensitivity analysis which is based on applying our MR workflow to external summary statistics. We focus on the hypothesis that increased heart failure risk can be seen as a consequence of changes in $PDSR_{rr}$.

Out of the 9304837 included in the diastolic function GWAS, 7064810 SNPs are also available in the published HERMES GWAS for heart failure. Clumping is performed as described above on the 7064810 candidate SNPs available in both GWAS. This leads to the selection of 17 SNPs for $PDSR_{rr}$ (instead of 20 if we consider our UK Biobank GWAS for heart failure). Supplementary Table 10 shows the results: the causal association between $PDSR_{rr}$ persists also if we use the GWAS results from the HERMES consortium. We note that effect sizes for the IVW method are also roughly comparable, strengthening our trust in the identified association.

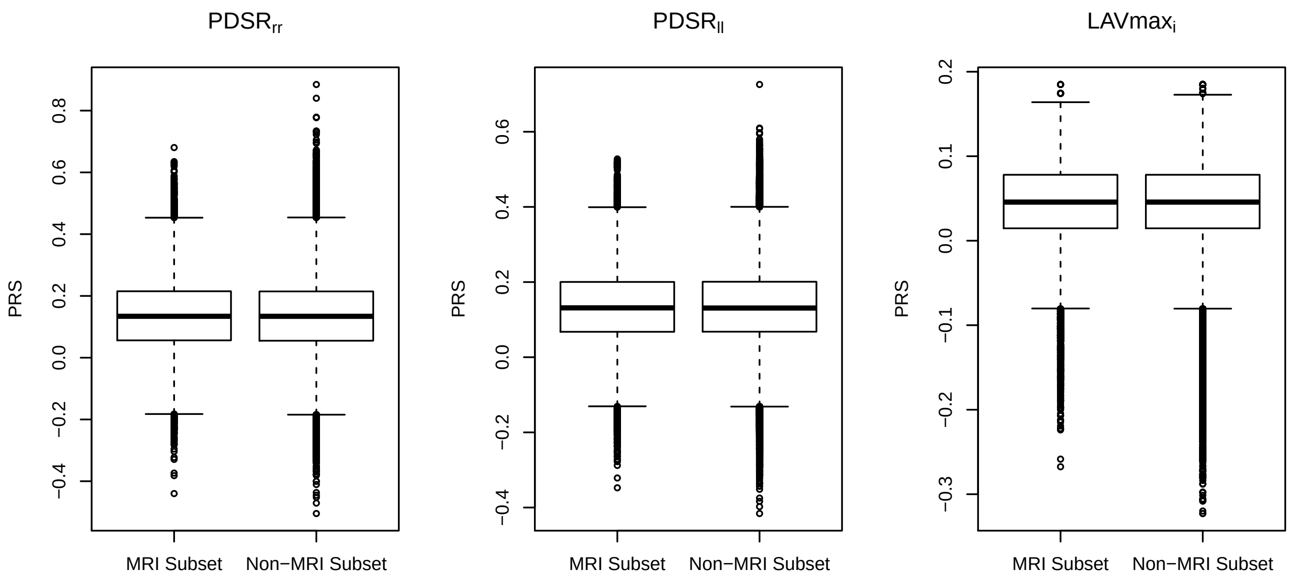


**Supplementary Figure 8.** Comparison of distribution of PRS within the MRI subset (subjects who were part of the first three releases of UKBB imaging data) vs. the non-MRI subset (subjects who were not part of the first three releases of UKBB imaging data)

**Supplementary Table 4.** Numerial results (parameter estimates/log(OR)) for PRS PheWAS analysis (Figure 5). PRS were scaled to represent 1 SD of diastolic function traits, quantitative traits were scaled to a SD of 1. SBP: systolic blood pressure, MAP: mean arterial pressure, DBP: diastolic blood pressure. Log: natural logarithm

| Phenotype | PRS for | Estimate | Lower CI | Upper CI | P-value |
|---|---|---|---|---|---|
| Alanine Aminotransferase (log) | $LAVmax_i$ | 0.08 | 0.02 | 0.13 | 1.15E-02 |
| | $PDSR_{rr}$ | -0.05 | -0.08 | -0.03 | 8.71E-05 |
| | $PDSR_{ll}$ | -0.02 | -0.05 | 0.01 | 2.50E-01 |
| Apolipoprotein b | $LAVmax_i$ | 0.07 | 0.00 | 0.13 | 3.93E-02 |
| | $PDSR_{rr}$ | -0.07 | -0.10 | -0.04 | 5.23E-07 |
| | $PDSR_{ll}$ | -0.03 | -0.06 | 0.00 | 8.27E-02 |
| BMI | $LAVmax_i$ | 0.10 | 0.04 | 0.16 | 1.21E-03 |
| | $PDSR_{rr}$ | 0.01 | -0.02 | 0.03 | 5.88E-01 |
| | $PDSR_{ll}$ | 0.01 | -0.02 | 0.04 | 6.97E-01 |
| Body water mass (whole body) | $LAVmax_i$ | -0.19 | -0.22 | -0.15 | 5.06E-27 |
| | $PDSR_{rr}$ | 0.02 | 0.01 | 0.03 | 6.44E-03 |
| | $PDSR_{ll}$ | 0.03 | 0.01 | 0.05 | 7.96E-04 |
| C-reactive protein (log) | $LAVmax_i$ | 0.09 | 0.03 | 0.15 | 5.40E-03 |
| | $PDSR_{rr}$ | -0.05 | -0.08 | -0.03 | 1.13E-04 |
| | $PDSR_{ll}$ | -0.04 | -0.07 | -0.01 | 1.67E-02 |
| Cardiomyopathy | $LAVmax_i$ | 0.39 | -0.41 | 1.19 | 3.39E-01 |
| | $PDSR_{rr}$ | -0.67 | -1.01 | -0.33 | 1.10E-04 |
| | $PDSR_{ll}$ | -0.02 | -0.42 | 0.38 | 9.21E-01 |
| Creatinine (log) | $LAVmax_i$ | 0.00 | -0.05 | 0.05 | 9.07E-01 |
| | $PDSR_{rr}$ | 0.02 | -0.00 | 0.04 | 6.00E-02 |
| | $PDSR_{ll}$ | 0.05 | 0.02 | 0.07 | 2.09E-04 |
| Cystatin C (log) | $LAVmax_i$ | 0.01 | -0.05 | 0.06 | 8.47E-01 |
| | $PDSR_{rr}$ | 0.06 | 0.03 | 0.08 | 2.70E-06 |
| | $PDSR_{ll}$ | 0.08 | 0.05 | 0.11 | 7.89E-08 |
| DBP (adjusted) | $LAVmax_i$ | 0.22 | 0.16 | 0.28 | 1.25E-12 |
| | $PDSR_{rr}$ | -0.08 | -0.11 | -0.06 | 6.52E-10 |
| | $PDSR_{ll}$ | -0.10 | -0.13 | -0.07 | 6.92E-11 |

P: P-value, not adjusted for multiplicity. Lower CI and upper CI refers to 95% confidence intervals.

**Supplementary Table 5.** Numerial results (parameter estimates/log(OR)) for PRS PheWAS analysis (Figure 5). PRS were scaled to represent 1 SD of diastolic function traits, quantitative traits were scaled to a SD of 1. SBP: systolic blood pressure, MAP: mean arterial pressure, DBP: diastolic blood pressure. Log: natural logarithm

| Phenotype | PRS for | Estimate | Lower CI | Upper CI | P-value |
|---|---|---|---|---|---|
| Dilated cardiomyopathy | $LAVmax_i$ | -0.48 | -1.55 | 0.58 | 3.75E-01 |
| | $PDSR_{rr}$ | -1.36 | -1.83 | -0.90 | 7.11E-09 |
| | $PDSR_{ll}$ | -0.85 | -1.39 | -0.31 | 1.94E-03 |
| eGFR (crea) | $LAVmax_i$ | -0.01 | -0.07 | 0.04 | 6.90E-01 |
| | $PDSR_{rr}$ | -0.02 | -0.05 | 0.00 | 5.40E-02 |
| | $PDSR_{ll}$ | -0.05 | -0.08 | -0.03 | 1.28E-04 |
| Fat free mass (whole body) | $LAVmax_i$ | -0.19 | -0.22 | -0.15 | 3.57E-27 |
| | $PDSR_{rr}$ | 0.02 | 0.01 | 0.03 | 7.29E-03 |
| | $PDSR_{ll}$ | 0.03 | 0.01 | 0.04 | 1.40E-03 |
| FEV1/FVC | $LAVmax_i$ | 0.15 | 0.09 | 0.22 | 7.89E-07 |
| | $PDSR_{rr}$ | -0.04 | -0.06 | -0.01 | 5.07E-03 |
| | $PDSR_{ll}$ | -0.06 | -0.09 | -0.03 | 1.05E-04 |
| Gamma-Glutamyltransferase (log) | $LAVmax_i$ | 0.10 | 0.04 | 0.16 | 1.33E-03 |
| | $PDSR_{rr}$ | -0.06 | -0.09 | -0.04 | 9.11E-07 |
| | $PDSR_{ll}$ | -0.05 | -0.08 | -0.02 | 5.96E-04 |
| HDL cholesterol | $LAVmax_i$ | -0.04 | -0.10 | 0.02 | 2.25E-01 |
| | $PDSR_{rr}$ | 0.04 | 0.02 | 0.07 | 1.15E-03 |
| | $PDSR_{ll}$ | -0.02 | -0.05 | 0.01 | 1.49E-01 |
| Heart failure | $LAVmax_i$ | 0.11 | -0.22 | 0.43 | 5.13E-01 |
| | $PDSR_{rr}$ | -0.30 | -0.44 | -0.16 | 2.09E-05 |
| | $PDSR_{ll}$ | -0.17 | -0.33 | -0.01 | 4.29E-02 |
| IGF-1 | $LAVmax_i$ | -0.04 | -0.10 | 0.02 | 1.57E-01 |
| | $PDSR_{rr}$ | -0.08 | -0.10 | -0.05 | 1.96E-09 |
| | $PDSR_{ll}$ | -0.04 | -0.07 | -0.01 | 1.06E-02 |
| Impedance (left arm) | $LAVmax_i$ | -0.08 | -0.12 | -0.04 | 1.70E-04 |
| | $PDSR_{rr}$ | -0.04 | -0.05 | -0.02 | 8.76E-05 |
| | $PDSR_{ll}$ | -0.03 | -0.06 | -0.01 | 1.30E-03 |
| Impedance (whole body) | $LAVmax_i$ | -0.06 | -0.11 | -0.02 | 9.29E-03 |
| | $PDSR_{rr}$ | -0.04 | -0.06 | -0.02 | 1.61E-04 |
| | $PDSR_{ll}$ | -0.03 | -0.05 | -0.00 | 3.03E-02 |
| LDL cholesterol | $LAVmax_i$ | 0.06 | -0.01 | 0.12 | 8.08E-02 |
| | $PDSR_{rr}$ | -0.05 | -0.08 | -0.03 | 1.08E-04 |
| | $PDSR_{ll}$ | -0.03 | -0.06 | 0.00 | 7.38E-02 |
| Leucocytes (count, log) | $LAVmax_i$ | 0.04 | -0.02 | 0.10 | 1.94E-01 |
| | $PDSR_{rr}$ | -0.09 | -0.12 | -0.07 | 1.43E-12 |
| | $PDSR_{ll}$ | -0.05 | -0.09 | -0.02 | 5.45E-04 |

P: P-value, not adjusted for multiplicity. Lower CI and upper CI refers to 95% confidence intervals.

**Supplementary Table 6.** Numerial results (parameter estimates/log(OR)) for PRS PheWAS analysis (Figure 5). PRS were scaled to represent 1 SD of diastolic function traits, quantitative traits were scaled to a SD of 1. SBP: systolic blood pressure, MAP: mean arterial pressure, DBP: diastolic blood pressure. Log: natural logarithm

| Phenotype | PRS for | Estimate | Lower CI | Upper CI | P |
|---|---|---|---|---|---|
| Mean platelet volumne | $LAVmax_i$ | 0.02 | -0.04 | 0.09 | 4.39E-01 |
| | $PDSR_{rr}$ | 0.01 | -0.02 | 0.04 | 4.42E-01 |
| | $PDSR_{ll}$ | 0.09 | 0.06 | 0.12 | 5.78E-09 |
| Platelet count | $LAVmax_i$ | 0.00 | -0.06 | 0.06 | 9.48E-01 |
| | $PDSR_{rr}$ | -0.05 | -0.07 | -0.02 | 2.37E-04 |
| | $PDSR_{ll}$ | -0.04 | -0.07 | -0.01 | 5.70E-03 |
| Platelet crit | $LAVmax_i$ | 0.01 | -0.04 | 0.07 | 6.40E-01 |
| | $PDSR_{rr}$ | -0.05 | -0.08 | -0.03 | 2.79E-05 |
| | $PDSR_{ll}$ | 0.00 | -0.03 | 0.03 | 9.09E-01 |
| Protein | $LAVmax_i$ | 0.05 | -0.01 | 0.12 | 1.18E-01 |
| | $PDSR_{rr}$ | -0.07 | -0.10 | -0.05 | 1.69E-07 |
| | $PDSR_{ll}$ | -0.05 | -0.09 | -0.02 | 9.40E-04 |
| Pulse rate | $LAVmax_i$ | -0.07 | -0.13 | -0.01 | 3.39E-02 |
| | $PDSR_{rr}$ | -0.30 | -0.33 | -0.28 | 5.86E-108 |
| | $PDSR_{ll}$ | -0.35 | -0.38 | -0.32 | 8.49E-103 |
| SBP (adjusted) | $LAVmax_i$ | 0.32 | 0.26 | 0.38 | 1.07E-27 |
| | $PDSR_{rr}$ | 0.02 | -0.00 | 0.05 | 6.08E-02 |
| | $PDSR_{ll}$ | 0.02 | -0.01 | 0.05 | 2.55E-01 |
| SHBG (log) | $LAVmax_i$ | -0.03 | -0.09 | 0.02 | 2.50E-01 |
| | $PDSR_{rr}$ | 0.09 | 0.06 | 0.11 | 8.20E-12 |
| | $PDSR_{ll}$ | -0.01 | -0.04 | 0.02 | 6.43E-01 |
| Standing height | $LAVmax_i$ | -0.50 | -0.54 | -0.46 | 2.31E-122 |
| | $PDSR_{rr}$ | 0.01 | -0.00 | 0.03 | 1.10E-01 |
| | $PDSR_{ll}$ | 0.04 | 0.02 | 0.07 | 3.11E-05 |
| Testosterone | $LAVmax_i$ | -0.01 | -0.04 | 0.02 | 6.13E-01 |
| | $PDSR_{rr}$ | 0.03 | 0.02 | 0.04 | 1.31E-05 |
| | $PDSR_{ll}$ | -0.00 | -0.02 | 0.01 | 8.90E-01 |
| Triglycerides (log) | $LAVmax_i$ | 0.05 | -0.02 | 0.11 | 1.41E-01 |
| | $PDSR_{rr}$ | -0.13 | -0.15 | -0.10 | 1.50E-21 |
| | $PDSR_{ll}$ | -0.03 | -0.06 | -0.00 | 4.08E-02 |

P: P-value, not adjusted for multiplicity. Lower CI and upper CI refers to 95% confidence intervals.

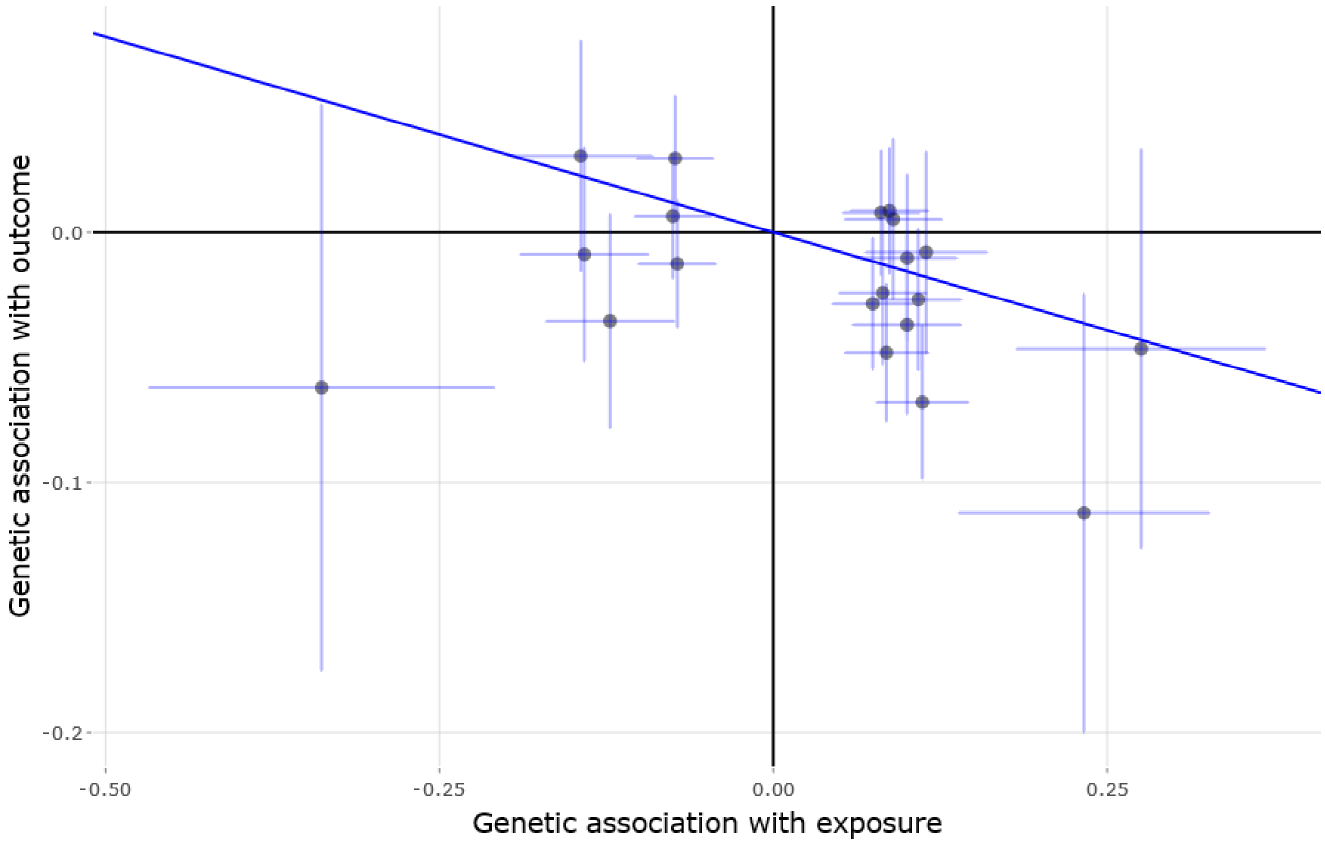


**Supplementary Figure 9. MR-Plot for the hypothesis that changes in PDSR$_{rr}$ (exposure) causes changes in heart failure risk (outcome).** Each dot represents one SNP, the lines represent confidence intervals. The regression line represents the IVW estimate.

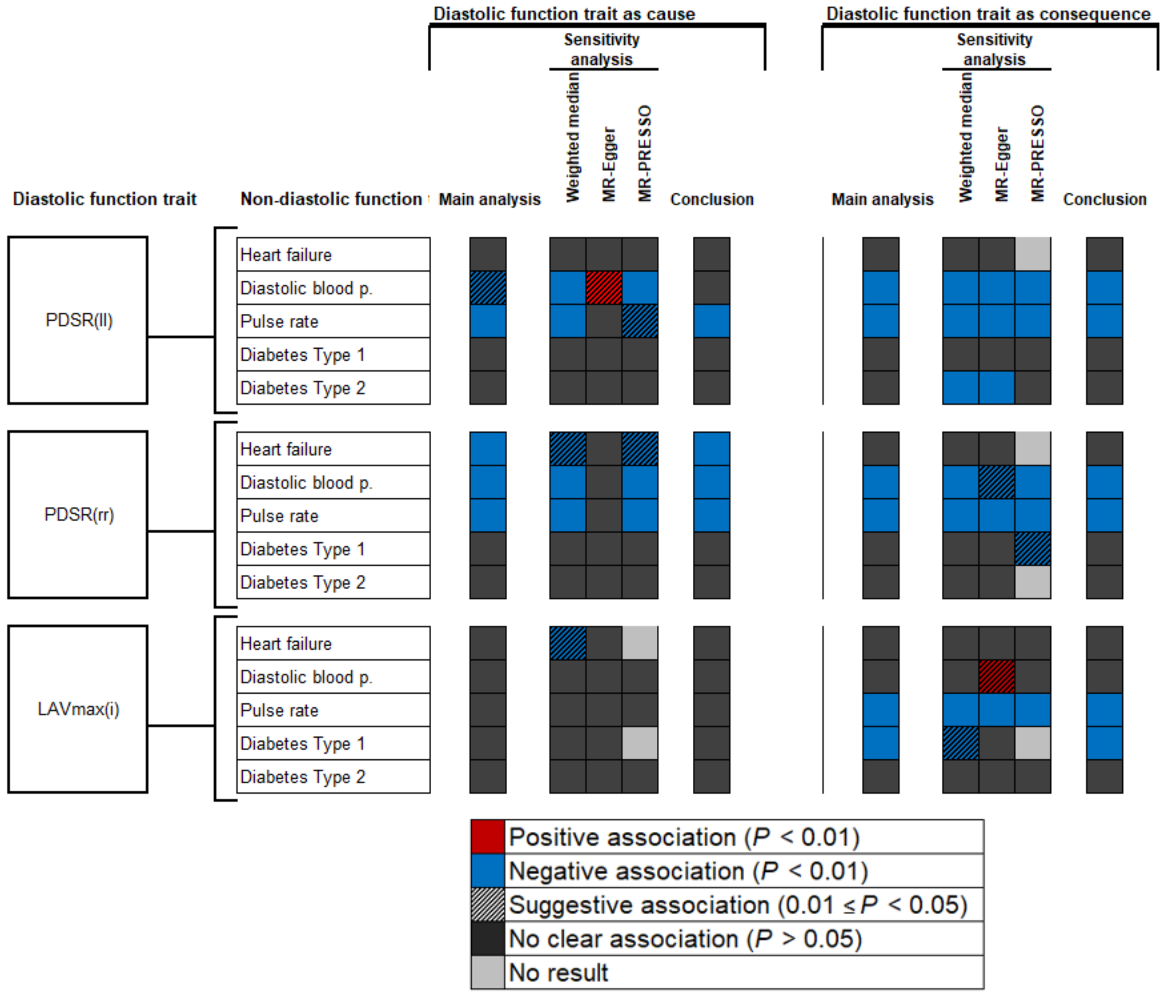


**Supplementary Figure 10. Summary of results from MR analyses of traits describing diastolic strain plus $LAVmax_i$ and multiple cardiac outcomes and phenotypes.** The style of this figure is adapted from van Oort et al.[87].“No result”, no outlier was detected with MR-PRESSO and therefore the MR-PRESSO estimate was identical to the IVW estimate. Detailed results are provided in Supplementary Tables 7, 8 and 9.

**Supplementary Table 7.** Numerial results (parameter estimates) for MR-experiments for PDSR$_{rr}$ vs. non-diastolic function traits (Figure 6). P-values are not adjusted for multiple testing. NA for MR-PRESSO means that no outlier were identified and therefore no result was calculated.

| Causal trait | Consequence trait | Method | Estimate | Sd | P |
|---|---|---|---|---|---|
| PDSR$_{rr}$ | Heart failure | Weighted median | -1.51E-01 | 6.12E-02 | 1.33E-02 |
| | | IVW | -1.56E-01 | 5.90E-02 | 8.04E-03 |
| | | MR-Egger | -1.03E-01 | 1.90E-01 | 5.88E-01 |
| | | MR-PRESSO | -1.23E-01 | 5.52E-02 | 3.90E-02 |
| PDSR$_{rr}$ | Diastolic blood pressure | Weighted median | -5.63E-01 | 1.25E-01 | 6.94E-06 |
| | | IVW | -5.03E-01 | 1.94E-01 | 9.35E-03 |
| | | MR-Egger | -5.42E-01 | 6.23E-01 | 3.84E-01 |
| | | MR-PRESSO | -4.67E-01 | 1.06E-01 | 4.95E-04 |
| PDSR$_{rr}$ | Pulse rate | Weighted median | -9.04E-01 | 1.82E-01 | 7.13E-07 |
| | | IVW | -1.74E+00 | 4.68E-01 | 2.08E-04 |
| | | MR-Egger | -2.17E+00 | 1.50E+00 | 1.49E-01 |
| | | MR-PRESSO | -1.01E+00 | 1.72E-01 | 1.54E-04 |
| PDSR$_{rr}$ | Diabetes T1 | Weighted median | 3.20E-02 | 1.03E-01 | 7.55E-01 |
| | | IVW | 2.33E-01 | 2.42E-01 | 3.35E-01 |
| | | MR-Egger | 2.03E-01 | 7.89E-01 | 7.96E-01 |
| | | MR-PRESSO | -2.36E-02 | 6.66E-02 | 7.28E-01 |
| PDSR$_{rr}$ | Diabetes T2 | Weighted median | -2.33E-02 | 4.13E-02 | 5.73E-01 |
| | | IVW | -4.73E-02 | 3.99E-02 | 2.36E-01 |
| | | MR-Egger | 1.33E-01 | 1.20E-01 | 2.68E-01 |
| | | MR-PRESSO | -6.73E-02 | 3.60E-02 | 7.80E-02 |
| Heart failure | PDSR$_{rr}$ | Weighted median | -7.35E-02 | 9.34E-02 | 4.31E-01 |
| | | IVW | -1.30E-02 | 8.64E-02 | 8.81E-01 |
| | | MR-Egger | 1.14E-03 | 2.24E-01 | 9.96E-01 |
| | | MR-PRESSO | NA | NA | NA |
| Diastolic blood pressure | PDSR$_{rr}$ | Weighted median | -1.85E-02 | 4.19E-03 | 1.07E-05 |
| | | IVW | -2.30E-02 | 3.01E-03 | 2.10E-14 |
| | | MR-Egger | -1.68E-02 | 8.34E-03 | 4.43E-02 |
| | | MR-PRESSO | -2.24E-02 | 2.94E-03 | 5.45E-14 |
| Pulse rate | PDSR$_{rr}$ | Weighted median | -9.19E-02 | 4.84E-03 | 2.22E-80 |
| | | IVW | -8.56E-02 | 3.40E-03 | 1.09E-139 |
| | | MR-Egger | -9.75E-02 | 7.32E-03 | 0.00E+00 |
| | | MR-PRESSO | -8.41E-02 | 3.28E-03 | 1.59E-108 |
| Diabetes T1 | PDSR$_{rr}$ | Weighted median | -1.89E-02 | 1.44E-02 | 1.89E-01 |
| | | IVW | -1.62E-02 | 1.16E-02 | 1.63E-01 |
| | | MR-Egger | -4.97E-02 | 2.59E-02 | 5.57E-02 |
| | | MR-PRESSO | -2.17E-02 | 1.09E-02 | 4.98E-02 |
| Diabetes T2 | PDSR$_{rr}$ | Weighted median | 2.57E-02 | 3.15E-02 | 4.15E-01 |
| | | IVW | -8.96E-03 | 1.97E-02 | 6.50E-01 |
| | | MR-Egger | -4.07E-02 | 4.82E-02 | 3.99E-01 |
| | | MR-PRESSO | NA | NA | NA |

P: P-value, not adjusted for multiplicity. Sd: Standard deviation, IVW: inverse-variance-weighted method

**Supplementary Table 8.** Numerical results (parameter estimates) for MR-experiments for $PDSR_{II}$ vs. non-diastolic function traits (Figure 6). P-values are not adjusted for multiple testing. NA for MR-PRESSO means that no outlier were identified and therefore no result was calculated.

| Causal trait | Consequence trait | Method | Estimate | Sd | P |
|---|---|---|---|---|---|
| $PDSR_{II}$ | Heart failure | Weighted median | 1.84E-01 | 2.12E-01 | 3.87E-01 |
| | | IVW | -4.19E-02 | 2.30E-01 | 8.56E-01 |
| | | MR-Egger | -6.54E-02 | 9.04E-01 | 9.42E-01 |
| | | MR-PRESSO | 1.11E-01 | 1.75E-01 | 5.37E-01 |
| $PDSR_{II}$ | Diastolic blood pressure | Weighted median | -1.74E+00 | 5.09E-01 | 6.34E-04 |
| | | IVW | -2.05E+00 | 8.13E-01 | 1.17E-02 |
| | | MR-Egger | 5.03E+00 | 2.50E+00 | 4.40E-02 |
| | | MR-PRESSO | -1.78E+00 | 4.74E-01 | 3.75E-03 |
| $PDSR_{II}$ | Pulse rate | Weighted median | -5.37E+00 | 8.33E-01 | 1.15E-10 |
| | | IVW | -6.45E+00 | 2.13E+00 | 2.51E-03 |
| | | MR-Egger | 1.04E+00 | 8.17E+00 | 8.99E-01 |
| | | MR-PRESSO | -5.23E+00 | 9.55E-01 | 1.20E-02 |
| $PDSR_{II}$ | Diabetes T1 | Weighted median | -5.37E-01 | 3.97E-01 | 1.76E-01 |
| | | IVW | 3.75E-01 | 9.13E-01 | 6.81E-01 |
| | | MR-Egger | 3.73E-02 | 3.64E+00 | 9.92E-01 |
| | | MR-PRESSO | -5.44E-01 | 2.97E-01 | 9.02E-02 |
| $PDSR_{II}$ | Diabetes T2 | Weighted median | -1.79E-01 | 1.55E-01 | 2.49E-01 |
| | | IVW | -1.03E-01 | 1.72E-01 | 5.49E-01 |
| | | MR-Egger | 3.45E-01 | 6.57E-01 | 5.99E-01 |
| | | MR-PRESSO | -1.19E-01 | 1.32E-01 | 3.88E-01 |
| Heart failure | $PDSR_{II}$ | Weighted median | -3.11E-02 | 2.87E-02 | 2.79E-01 |
| | | IVW | -1.11E-02 | 2.66E-02 | 6.78E-01 |
| | | MR-Egger | 2.56E-03 | 6.91E-02 | 9.70E-01 |
| | | MR-PRESSO | NA | NA | NA |
| Diastolic blood pressure | $PDSR_{II}$ | Weighted median | -8.92E-03 | 1.22E-03 | 2.38E-13 |
| | | IVW | -1.04E-02 | 8.44E-04 | 8.39E-35 |
| | | MR-Egger | -8.21E-03 | 2.34E-03 | 4.41E-04 |
| | | MR-PRESSO | -1.02E-02 | 8.21E-04 | 1.08E-33 |
| Pulse rate | $PDSR_{II}$ | Weighted median | -2.57E-02 | 1.44E-03 | 9.90E-71 |
| | | IVW | -2.56E-02 | 9.92E-04 | 3.68E-147 |
| | | MR-Egger | -2.76E-02 | 2.14E-03 | 0.00E+00 |
| | | MR-PRESSO | -2.53E-02 | 9.62E-04 | 3.96E-112 |
| Diabetes T1 | $PDSR_{II}$ | Weighted median | -5.62E-03 | 4.11E-03 | 1.72E-01 |
| | | IVW | -4.81E-04 | 2.99E-03 | 8.72E-01 |
| | | MR-Egger | -6.40E-03 | 6.72E-03 | 3.41E-01 |
| | | MR-PRESSO | -2.16E-03 | 2.67E-03 | 4.21E-01 |
| Diabetes T2 | $PDSR_{II}$ | Weighted median | -2.33E-02 | 8.39E-03 | 5.50E-03 |
| | | IVW | -6.69E-03 | 5.72E-03 | 2.42E-01 |
| | | MR-Egger | -3.82E-02 | 1.38E-02 | 5.53E-03 |
| | | MR-PRESSO | -8.37E-03 | 5.61E-03 | 1.37E-01 |

P: P-value, not adjusted for multiplicity. Sd: Standard deviation, IVW: inverse-variance-weighted method

**Supplementary Table 9.** Numerial results (parameter estimates) for MR-experiments for LAVmax$_i$ vs. non-diastolic function traits (Figure 6). P-values are not adjusted for multiple testing. NA for MR-PRESSO means that no outlier were identified and therefore no result was calculated.

| Causal trait | Consequence trait | Method | Estimate | Sd | P |
|---|---|---|---|---|---|
| LAVmax$_i$ | Heart failure | Weighted median | -1.47E+00 | 6.66E-01 | 2.71E-02 |
| | | IVW | -1.30E+00 | 5.06E-01 | 1.02E-02 |
| | | MR-Egger | -1.53E-02 | 1.52E+00 | 9.92E-01 |
| | | MR-PRESSO | NA | NA | NA |
| LAVmax$_i$ | Diastolic blood pressure | Weighted median | 5.73E-01 | 1.44E+00 | 6.90E-01 |
| | | IVW | 4.90E+00 | 5.34E+00 | 3.58E-01 |
| | | MR-Egger | 4.69E+00 | 1.74E+01 | 7.87E-01 |
| | | MR-PRESSO | 6.21E-01 | 1.14E+00 | 6.08E-01 |
| LAVmax$_i$ | Pulse rate | Weighted median | -2.07E+00 | 1.44E+00 | 1.51E-01 |
| | | IVW | -2.81E+00 | 1.70E+00 | 9.75E-02 |
| | | MR-Egger | 1.21E+00 | 5.24E+00 | 8.17E-01 |
| | | MR-PRESSO | -1.54E+00 | 1.19E+00 | 2.41E-01 |
| LAVmax$_i$ | Diabetes T1 | Weighted median | -1.55E+00 | 1.34E+00 | 2.50E-01 |
| | | IVW | -5.81E-01 | 1.29E+00 | 6.52E-01 |
| | | MR-Egger | 1.36E-01 | 4.27E+00 | 9.75E-01 |
| | | MR-PRESSO | NA | NA | NA |
| LAVmax$_i$ | Diabetes T2 | Weighted median | -2.10E-01 | 4.82E-01 | 6.63E-01 |
| | | IVW | 4.72E-01 | 5.80E-01 | 4.16E-01 |
| | | MR-Egger | -7.19E-01 | 1.80E+00 | 6.90E-01 |
| | | MR-PRESSO | -4.76E-02 | 2.85E-01 | 8.73E-01 |
| Heart failure | LAVmax$_i$ | Weighted median | 2.31E-03 | 1.07E-02 | 8.29E-01 |
| | | IVW | 3.32E-03 | 1.23E-02 | 7.87E-01 |
| | | MR-Egger | 2.29E-02 | 3.13E-02 | 4.65E-01 |
| | | MR-PRESSO | -2.87E-03 | 7.58E-03 | 7.13E-01 |
| Diastolic blood pressure | LAVmax$_i$ | Weighted median | 6.53E-04 | 4.87E-04 | 1.80E-01 |
| | | IVW | 7.21E-04 | 3.71E-04 | 5.18E-02 |
| | | MR-Egger | 2.38E-03 | 1.03E-03 | 2.00E-02 |
| | | MR-PRESSO | 5.16E-04 | 3.60E-04 | 1.53E-01 |
| Pulse rate | LAVmax$_i$ | Weighted median | -4.58E-03 | 5.42E-04 | 3.13E-17 |
| | | IVW | -4.23E-03 | 3.80E-04 | 9.46E-29 |
| | | MR-Egger | -5.27E-03 | 8.18E-04 | 1.23E-10 |
| | | MR-PRESSO | -4.35E-03 | 3.70E-04 | 9.86E-30 |
| Diabetes T1 | LAVmax$_i$ | Weighted median | -3.03E-03 | 1.50E-03 | 4.29E-02 |
| | | IVW | -3.28E-03 | 1.14E-03 | 4.09E-03 |
| | | MR-Egger | -2.44E-03 | 2.58E-03 | 3.43E-01 |
| | | MR-PRESSO | NA | NA | NA |
| Diabetes T2 | LAVmax$_i$ | Weighted median | 3.28E-03 | 3.40E-03 | 3.35E-01 |
| | | IVW | -9.60E-04 | 2.41E-03 | 6.90E-01 |
| | | MR-Egger | 3.64E-03 | 5.87E-03 | 5.36E-01 |
| | | MR-PRESSO | -7.81E-04 | 2.30E-03 | 7.34E-01 |

P: P-value, not adjusted for multiplicity. Sd: Standard deviation, IVW: inverse-variance-weighted method

**Supplementary Table 10.** Overview on MR results obtained with HERMES consortium as heart failure GWAS: assessment of changes in $PDSR_{rr}$ as causes for heart failure. No multiplicity adjustment is performed.

| Trait | Estimate | Sd | P |
|---|---|---|---|
| IVW | -0.1456 | 0.0543 | 0.0073 |
| Weighted median | -0.1080 | 0.0480 | 0.0137 |
| MR-Egger | 0.1178 | 0.1820 | 0.5176 |
| MR-PRESSO | -0.7139 | 0.0332 | 0.0529 |

P: P-value, not adjusted for multiplicity. Sd: Standard deviation, IVW: inverse-variance-weighted method
